# Detection of enterovirus RNA in pancreas and lymphoid tissues of organ donors with type 1 diabetes

**DOI:** 10.1101/2024.09.11.24313112

**Authors:** Jutta E Laiho, Sami Oikarinen, Sofia Morfopoulou, Maarit Oikarinen, Ashlie Renner, Daniel Depledge, Matthew C Ross, Ivan C Gerling, Judith Breuer, Joseph F Petrosino, Vincent Plagnol, Alberto Pugliese, Antonio Toniolo, Richard E Lloyd, Heikki Hyöty, JDRF nPOD-Virus Group

**Affiliations:** Department of Virology, Faculty of Medicine and Health Technology, Tampere University, Finland; Department of Infection, Immunity and Inflammation, UCL Great Ormond Street Institute of Child Health, University College London, London, UK; Department of Molecular Virology and Microbiology, Baylor College of Medicine, Houston, TX 77030, USA; NYU, Grossman School of Medicine, New York, New York, United States; Alkek Center for Metagenomics and Microbiome Research, Baylor College of Medicine, Houston, TX 77030, USA; Division of Endocrinology, Diabetes, and Metabolism, Department of Medicine, University of Tennessee Health Science Center, Memphis, TN, United States; Genomics PLC, Oxford, Oxfordshire, UK; Department of Diabetes Immunology, Arthur Riggs Diabetes & Metabolism Research Institute, Beckmann Research Institute, City of Hope, Duarte, United States; Global Virus Network, University of Insubria, Varese, Italy; Fimlab Laboratories, Tampere, Finland; Department of Pediatrics, Tampere University Hospital, Tampere, Finland

**Author notes:** Joint first authors (Jutta E Laiho, Sami Oikarinen). Corresponding author: Jutta E Laiho, Department of Virology, Faculty of Medicine and Health Technology, Tampere University, Finland.

**Keywords:** Type 1 diabetes, Enterovirus, Pancreas, Lymph nodes, Spleen, Persistent infection, Islet autoimmunity, Organ donors

## Abstract

**Aims/hypothesis:** The nPOD-Virus group collaboratively applied innovative technologies to detect and sequence viral RNA in pancreas and other tissues from organ donors with type 1 diabetes. These analyses involved the largest number of pancreas samples collected to date.

**Methods:** We analysed pancreas, spleen, pancreatic lymph nodes, and duodenum samples from the following donor groups: a) donors with type 1 diabetes (n=71), with (n=35) or without (n=36) insulin-containing islets, (b) donors with single or double islet autoantibody positivity without diabetes (n=22) and c) autoantibody-negative donors without diabetes (control donors) (n=74). Five research laboratories participated in this collaborative effort using approaches for unbiased discovery of RNA viruses (two RNA-Seq platforms), targeted detection of *Enterovirus A-D* species using RT-PCR, and tests for virus growth in cell-culture.

**Results:** Direct RNA-Seq did not detect virus signal in pancreas samples, whereas RT-PCR detected enterovirus RNA confirmed by sequencing in low amounts in pancreas samples in three of the five donor groups, namely donors with type 1 diabetes with insulin-containing islets, 16% (5/32) donors being positive, donors with single islet autoantibody positivity with 53% (8/15) donors being positive, and non-diabetic donors with 8% (4/49) being enterovirus RNA positive. Detection of enterovirus RNA was significantly more frequent in single islet autoantibody-positive donors compared to donors with type 1 diabetes with insulin-deficient islets (p-value <0.001) and control donors (p-value 0.004). In some donors, pancreatic lymph nodes were also positive. RT-PCR detected enterovirus RNA also in spleen of a small number of donors and virus enrichment in susceptible cell lines before RT-PCR resulted in much higher rate in spleen positivity, particularly in donors with type 1 diabetes. Interestingly, the enterovirus strains detected did not cause a typical lytic infection, possibly reflecting their persistence-prone nature.

**Conclusions/interpretation:** This was the largest coordinated effort to examine the presence of enterovirus RNA in pancreas of organ donors with type 1 diabetes, using a multitude of assays. These findings are consistent with the notion that both the subjects with type 1 diabetes and those with islet autoantibodies may carry a low-grade enterovirus infection in the pancreas and lymphoid tissues.

**Research in context:** What is already known about this subject?

- Enterovirus infections are among the prime candidates for environmental triggers of type 1 diabetes.
- Pancreas (and other tissue) samples of subjects with type 1 diabetes have not been extensively studied for the presence of enterovirus RNA.

What is the key question?

- Can enterovirus RNA be detected in the pancreas and lymphoid tissues of individuals with and without type 1 diabetes?

What are the new findings?

- Enterovirus RNA can be detected in low amounts in the pancreas and lymphoid tissues using selected enterovirus-specific methods.
- Detection of enterovirus RNA in the pancreas was most frequent in prediabetic subjects.
- Enterovirus RNA was found also in pancreatic lymph nodes and in spleen where it was more frequently detected in donors with type 1 diabetes compared to non-diabetic donors, with properties previously observed in persistent infections.

How might this impact on clinical practice in the foreseeable future?

- The findings support the enterovirus - type 1 diabetes association and may have an effect on the primary and secondary prevention strategies towards the disease.

## Introduction

The presence of enterovirus VP1 capsid protein in pancreatic islets (1–3), mostly in beta cells, has been associated with type 1 diabetes; some studies have also found enterovirus RNA in the islets (3–6). Studies of infants who passed from acute enterovirus infections demonstrated virus in pancreatic islets, causing inflammation and cell damage (7,8). The DiViD study, which examined pancreatic tissue obtained at biopsy from 6 adult newly diagnosed patients found evidence of enterovirus infection in the islets of all patients (9). Thus, enteroviruses have tropism for pancreatic islet cells, including beta cells, which is also observed in cell models where enterovirus readily infect human beta cells and impair insulin secretion (10). Such tropism to beta cells could be explained by the abundant expression of coxsackie and adenovirus receptor (CAR) (11), which is used by the coxsackie B group enteroviruses linked to type 1 diabetes (12). The reduced ability of beta cells to generate innate immune responses may contribute to their susceptibility to viral infection (13), and perhaps limit ability to fully clear the infection.

Previous studies suggested that enteroviruses can undergo terminal deletion in myocardium and pancreas, becoming replication-defective and favouring persistence as a low-grade chronic infection (14,15) which may be less susceptible to immune clearance. Prolonged, persisting infection of replication defective viruses could explain the high detection rate of enterovirus protein or RNA in the pancreatic islets of patients with type 1 diabetes (80%-100% of patients) and the challenge of isolating replication-competent virus from their pancreas (9). In addition, only very few beta cells are positive for enterovirus protein and the levels of viral RNA are extremely low in the pancreatic islets. This does not fit with a typical acute phase of infection where usually several cells are infected producing large amounts of virus. In addition to the pancreatic islets of patients with type 1 diabetes, some studies have detected enteroviruses in the small intestinal mucosa, a well-known EV replication site (16–18).

The JDRF nPOD (Network for Pancreatic Organ Donor with Diabetes) has collected pancreas and other tissue specimens from organ donors with type 1 diabetes through a wide spectrum of age and disease duration. The nPOD-Virus Group has been established as an international collaboration to investigate viral infections in pancreas and other tissues. Here we report the group efforts focused on detecting and sequencing viral RNA in pancreas, spleen, pancreatic lymph nodes (PLN), and duodenum. Independent laboratories in Finland, Italy, the U.S.A. and the U.K., developed and applied, for the first time, RNA-Seq methods for the unbiased discovery of any RNA virus potentially present in the pancreas of type 1 diabetic patients. Based on the pre-existing association data with enteroviruses, the group also deployed highly sensitive, enterovirus-specific detection methods to directly address the presence of enterovirus RNA.

## Research design and methods

### Organ donors and tissues

We examined tissue samples from cadaveric organ donors collected by nPOD. As part of the coordinated efforts of the nPOD-Virus Group, we investigated tissues from 167 organ donors: 71 donors with type 1 diabetes, of which 35 had residual insulin containing islets (T1D-ICI) and 36 only had insulin-deficient islets (T1D-IDI); 22 islet autoantibody (AAb) positive donors without diabetes considered at increased risk for type 1 diabetes, of whom 15 donors expressed a single autoantibody (AAb+), and 7 donors had ≥2 autoantibodies (AAb++). Finally, 74 autoantibody-negative donors without diabetes were included as a control group (ND). Demographic information for each group is summarized in **Table 1**. Detailed donor information is provided in **ESM Table 1**. Examined tissues included frozen pancreas samples from the majority of organ donors. From select cases, other organs were recovered: spleen, pancreatic lymph nodes, live cryopreserved lymphoid cells, duodenal mucosa. Samples were stored at −80°C. The standardised collection protocol for the tissues analysed is described in Campbell-Thompson et al (19). All samples were de-identified and obtained by nPOD through its partnership organ procurement organizations, after consent for organ donation and research was obtained from family members and the Health Center Review Board, University of Florida.

**Table 1.**
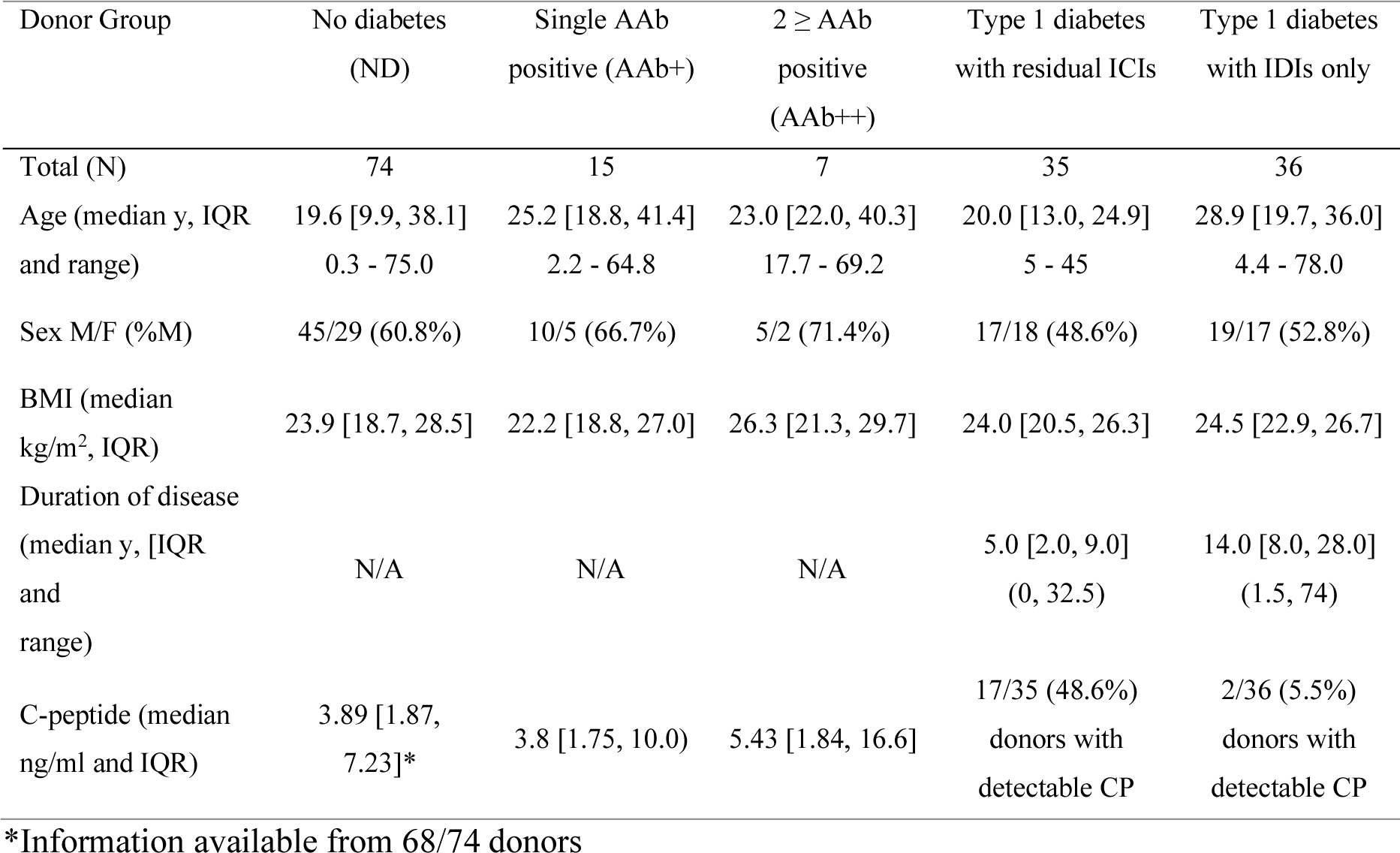
Donor demographics for each of the five donor groups. AAb, autoantibody; ICI, insulin containing islet; IDI, insulin deficient islet; N/A, not applicable; CP, C-peptide.

| Donor Group | No diabetes<br>(ND) | Single AAb<br>positive (AAb+) | 2 ≥ AAb<br>positive<br>(AAb++) | Type 1 diabetes<br>with residual ICIs | Type 1 diabetes<br>with IDIs only |
| --- | --- | --- | --- | --- | --- |
| Total (N) | 74 | 15 | 7 | 35 | 36 |
| Age (median y, IQR<br>and range) | 19.6 [9.9, 38.1]<br>0.3 - 75.0 | 25.2 [18.8, 41.4]<br>2.2 - 64.8 | 23.0 [22.0, 40.3]<br>17.7 - 69.2 | 20.0 [13.0, 24.9]<br>5 - 45 | 28.9 [19.7, 36.0]<br>4.4 - 78.0 |
| Sex M/F (%M) | 45/29 (60.8%) | 10/5 (66.7%) | 5/2 (71.4%) | 17/18 (48.6%) | 19/17 (52.8%) |
| BMI (median<br>kg/m<sup>2</sup>, IQR) | 23.9 [18.7, 28.5] | 22.2 [18.8, 27.0] | 26.3 [21.3, 29.7] | 24.0 [20.5, 26.3] | 24.5 [22.9, 26.7] |
| Duration of disease<br>(median y, [IQR<br>and<br>range) | N/A | N/A | N/A | 5.0 [2.0, 9.0]<br>(0, 32.5) | 14.0 [8.0, 28.0]<br>(1.5, 74) |
| C-peptide (median<br>ng/ml and IQR) | 3.89 [1.87,<br>7.23]* | 3.8 [1.75, 10.0] | 5.43 [1.84, 16.6] | 17/35 (48.6%)<br>donors with<br>detectable CP | 2/36 (5.5%)<br>donors with<br>detectable CP |

\*Information available from 68/74 donors

Five different laboratories performed independent assays using diverse methodologies to detect traces of enteroviruses or other microbes in pancreas and other tissues. A goal of the nPOD-Virus group was to approach the question about viral aetiology of type 1 diabetes and explore what type of viruses may be present, and if so, potentially associated with disease. To this end, we implemented two unbiased discovery approaches for microbes, based on two different RNA-Seq methods. In addition, based on pre-existing evidence of an association of type 1 diabetes with enterovirus infections, we employed enterovirus specific RT-PCR assays, and enterovirus propagation in cell cultures, followed by RT-PCR as well as enterovirus capsid protein staining. Samples from the donors were distributed to the five participating laboratories according to the protocol shown in **Fig. 1**.

**Figure 1.**
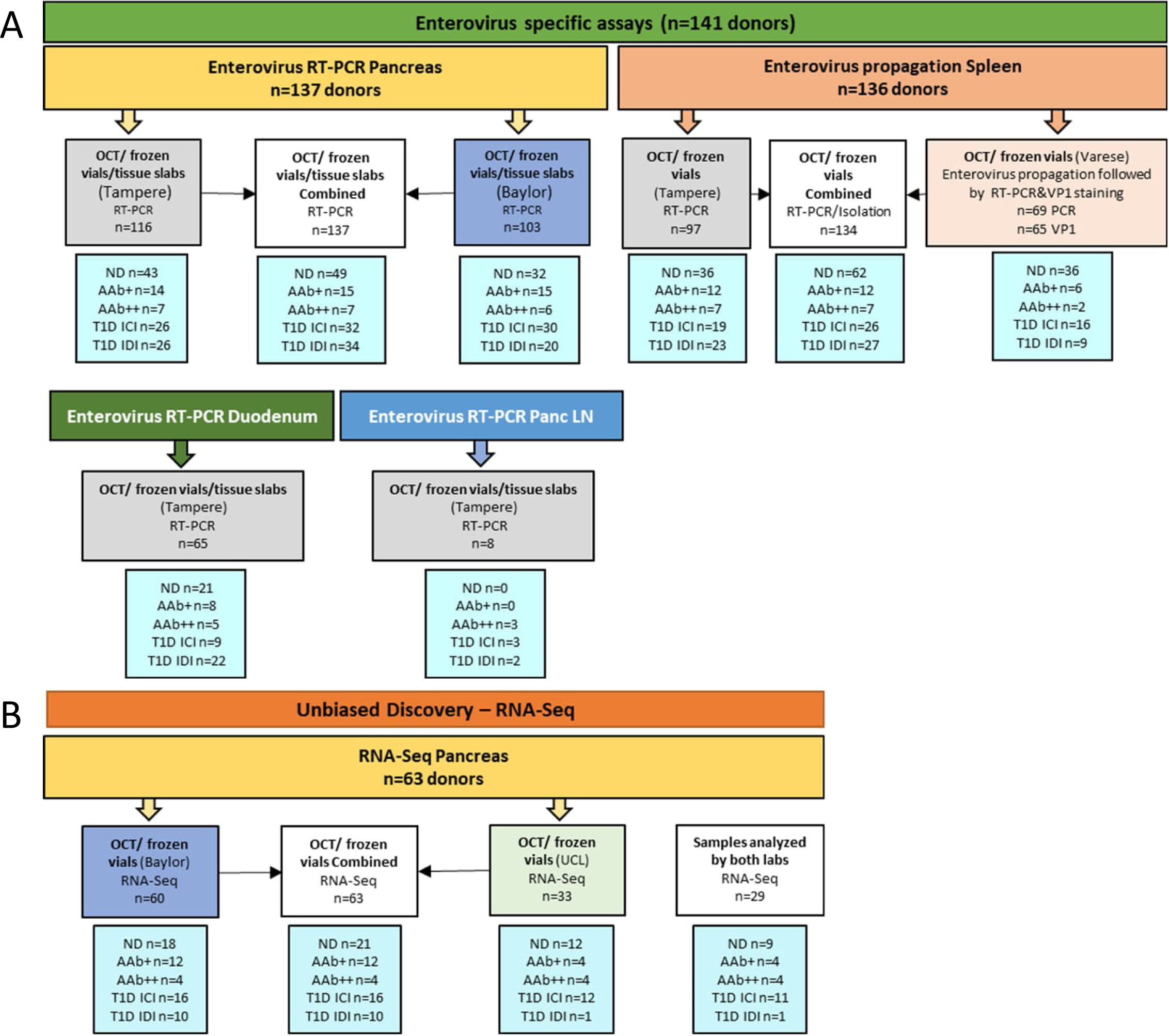
Study design and distribution of samples for analyses. (A) describes enterovirus specific assays and (B) describes the unbiased discovery (RNA-seq) showing the number of donors analysed in different laboratories. Two laboratories carried out enterovirus specific RT-PCR (Baylor and Tampere) and one lab (Varese) enterovirus propagation in cell cultures (A). The donors analysed in both labs altogether are marked as combined (white boxes). Similarly, two laboratories carried out RNA-Seq (Baylor and UCL) (B). Sample allocation depended on the assay performed, however, for each assay samples were distributed in various batches. Samples were either snap frozen tissue pieces, live lymphoid cells, or tissue pieces frozen in optimal cutting temperature (OCT) compound.

### Unbiased discovery of microbes

RNA-Seq studies were performed on pancreas samples in two laboratories at the University College London (UCL), London, UK, and at the Baylor College of Medicine (BCM), Houston, USA. Based on sample availability for coordinated studies, RNA-Seq analyses were performed on pancreas samples from 63 nPOD donors: 6 T1D-ICI, 10 T1D-IDI, 4 AAb++, 12 AAb+ and 21 ND donors. Of the above 63 donors, 29 were analysed in both laboratories (11 T1D-ICI, 1 T1D-IDI, 4 AAb++, 4 AAb+, 9 ND).

#### RNA-Seq analyses at UCL

Over four years, UCL sequenced frozen pancreas samples from 33 nPOD cases (12 T1D-ICI, 1 T1D-IDI, 4 AAb+, 4 AAb++ and 12 ND). We developed the methodology in four steps (described below) and used several extraction and library preparation approaches, to maximize sensitivity. Initial negative results motivated the development of a specific sequence capture method (20) to enrich for enteroviral sequences and adding the analysis of laser captured islet RNA to further increase sensitivity (21). The RNA obtained was then subjected to Illumina high throughput RNA sequencing.

*In step I* (first pilot stage), we examined pancreas from 3 T1D-ICI, 1 T1D-IDI and 2 ND cases, based on availability of optimal samples for RNA-Seq. Disease duration ranged from 4 to 28 years. *Step II* investigated tissues from donors with shorter disease duration to minimize the time between sample collection and T1D onset [4 T1D-ICI cases (disease duration range: 1-5 years), 4 AAb++ and 3 AAb+ cases]. In *step III* we examined pancreas from 4 T1D-ICI cases with enterovirus VP1 immuno-positivity by immunohistochemistry and HLA class I hyperexpression, along with 5 ND donors (for details, see immunohistochemistry results in the accompanying publication Rodriguez-Calvo et al, submitted in *Diabetologia*). From one donor with type 1 diabetes, two samples were analysed. Finally, in *step IV* we examined laser micro-dissected islets from 6 T1D-ICI, 4 autoantibody-positive (3 AAb++, 1 AAb+) and 6 ND donors. From one donor with type 1 diabetes, two samples were analysed.

In *steps I and II*, total RNA was isolated using Illumina GAIIx (step I) or the Illumina HiSeq2500 (step II), followed by a poly(A) selection step for mRNA. In *steps III and IV* we used the Agilent SureSelect system to enrich the potential enteroviral sequences in pancreatic samples. RNA extraction was performed as described (22). For double-stranded (ds) cDNA generation, we used a protocol optimized for RNA viruses (20,23). The ds-cDNA was sheared, and libraries prepared as per the SureSelect protocol v1.4. Enrichment for enteroviral sequences was performed using a set of 120-mer biotinylated RNA oligonucleotides prior to indexing and sequencing on different Illumina platforms (MiSeq, HiSeq, NextSeq). The bait set (RNA oligonucleotides) was designed using an in-house script written for an EU-funded project aimed at using SureSelect in a pathogen diagnostic setting (PathSeek). The bait set hybridized against all members of the *Enterovirus A* species (n=363 probes), *B* species (n=176) and *C* species (n=303), based on sequences available in Genbank at the time of design (15 May 2013). Up to 8 mismatches in a 120-mer oligo was accepted to still enable capture of the targeted sequence, ensuring enterovirus detection provided these shared a reasonable degree of similarity.

#### Positive control experiment for sequence capture

As positive control, ULC sequenced pancreatic tissue samples that were spiked in at different dilutions (10^-4 to 10^-8 range) of coxsackievirus B1 (CVB1) and a negative control, to assess the sensitivity of the sequence capture method prior to its use (**ESM data 1)**.

#### Metagenomic whole genome shotgun sequencing at BCM

BCM performed metagenomic whole genome shotgun (WGS) sequencing from 60 nPOD frozen pancreas samples (16 T1D-ICI, 10 T1D-IDI, 12 AAb+, 4 AAb++ and 18 ND). Total pancreatic nucleic acids were extracted using the MagMax Viral RNA Isolation Kit (Cat # AM1939, Thermo Fisher, Waltham, MA), without DNAse to prevent DNA removal. Extracted viral RNA was reverse transcribed using SuperScript II RT (Cat # 18064014, Thermo Fisher) and random hexamers. After short molecule and random hexamer removal with ChargeSwitch (Cat # CS12000, Thermo Fisher), molecules were amplified and tagged with a 12 base-pair barcode tag containing a V8A2 semi-random primer (BC12-V8A2 construct using AccuPrimeTM Taq polymerase and cleaned with ChargeSwitch kit). Tags were attached via a barcoded, semi-random primer construct resulting in dual barcoded (same barcode on both sides) amplified fragments. The indexes used were 12 bp Golay Barcodes. Separate negative controls were introduced during extraction, amplification, and library preparation steps. We performed a single WGS library prep per sequencing lane (without shearing) of pooled, pre-barcoded samples to minimize carry-over, as each lane only had a single index. Since all samples carried secondary internal barcodes, they were not subject to carry-over or cross-bleed that sometimes is observed from run to run with library indexes using the Illumina platform. The size of the library was verified via bioanalyzer to ensure appropriate range for the platform (∼200-1000 bp). The library was then loaded in an Illumina HiSeq2000 (Illumina, Carlsbad, CA) and sequenced using the 2×100bp chemistry at the Human Genome Sequencing Center, BCM. Reads were demultiplexed into a sample bin using the barcode prefixing read-1 and read-2, allowing zero mismatches. Demultiplexed reads were further processed by trimming off barcodes, semi-random primer sequences, and Illumina adapters. This process utilized a custom demultiplexer and the BBDuk algorithm included in BBMap53.

#### Bioinformatics analysis and community profiling at UCL and BCM

A dual analytic approach was implemented. Data was first analysed in unbiased manner, assuming no prior knowledge of potential pathogens and characterizing the full species profile for each sample. In addition, data was specifically searched for enteroviral sequences. PCR duplicates were removed with an in-house script that collapses read pairs by sequence identity using 90% of the sequence as signature. We removed low quality and low complexity sequences with PrinSeq (24) and human sequences with Novoalign (version V2.07.13 - human reference genome GRCh37) followed by BLASTn (25). High quality contigs of at least 200bp length were *de novo* assembled with Velvet (26). Contigs and the unassembled reads were annotated with BLASTx (default parameters) and a custom protein database consisting of viral, human microbiome bacterial, human and mouse RefSeq proteins (October 2013 version).

Coxsackievirus proteins that were not present in the RefSeq collection were added to the database. To search specifically for enterovirus sequences, we aligned (Novoalign V2.07.13) quality-controlled reads simultaneously to the genomes of enteroviruses from species A, B and D (NC_001612, NC_001472, NC_001430). This search was repeated using all enterovirus full genomes from GenBank (221 genomes, January 2020) and Bowtie2 (27). We employed metaMix 0.1 (28), which is used in clinical diagnostics for pathogen detection in brain biopsies from patients with encephalitis of unknown cause (29–34), to characterize the species that are present in each sample. The read support parameter cutoff for a species to be retained in the profile, was ten reads.

### Targeted enterovirus detection by RT-PCR

The presence of enterovirus RNA was assessed in tissues from 141 nPOD organ donors using a sensitive RT-PCR assay. Frozen pancreas samples from 137 nPOD organ donors (32 T1D-ICI, 34 T1D-IDI, 7 AAb++, 15 AAb+, 49 ND) were analysed in two laboratories at Tampere University, Finland, and in the Department of Molecular Virology and Microbiology, BCM, Houston, TX, USA. Based on sample availability, the Tampere laboratory also examined frozen spleen samples from 97 organ donors (19 T1D-ICI, 23 T1D-IDI, 7 AAb++, 12 AAb+ and 36 ND), pancreatic lymph node (PLN) samples from 8 organ donors (3 T1D-ICI, 2 T1D-IDI and 3 AAb++), and duodenum samples from 65 organ donors (9 T1D-ICI, 22 T1D-IDI, 5 AAb++, 8 AAb+ and 21 ND).

In Tampere, RNA was extracted from frozen tissue using the Viral RNA Kit (Qiagen, Hilden, Germany) and samples were analysed with a quantitative real-time RT-PCR method (35). In the BCM laboratory, pancreatic RNA was extracted with the MagMax Viral RNA Isolation Kit (Invitrogen; Thermo Fischer). RNA was converted to cDNA with Superscript III RT (Invitrogen) according to the manufacturer’s directions, with random primers. PCR was carried out with SYBR-Green PCR master mix (Invitrogen) using the same primers as in Tampere (35). PCR included a denaturation step (95 °C for 10 min) followed by 50 cycles of 95 °C for 30 s and 60 °C for 60 s. In both laboratories, positive RT-PCR signals were confirmed by sequencing the PCR amplicon and samples were considered positive only if an enterovirus sequence was obtained.

The degree of RNA degradation was analysed in selected pancreas, spleen and duodenum samples, using Agilent Fragment Analyzer.

### Enterovirus propagation in cell culture prior to RNA detection by RT-PCR

Enterovirus propagation in cell cultures was carried out for spleen samples at the University of Insubria, Varese, Italy, to amplify the virus prior to RT-PCR assays and immunostaining. For this approach we selected spleen samples since they do not contain enzymes that can affect cultured cells. Donors were selected according to the availability of live spleen cell suspensions for both controls and T1D donors. Snap frozen spleen tissue was also tested in the form of spleen homogenates. We could examine samples from 69 donors (16 T1D-ICI, 9 T1D-IDI, 2 AAb++, 6 AAb+, 36 ND). A published procedure for detecting persistent enterovirus infections (9,36) was followed with minor modifications. Briefly, to enrich for virus nPOD spleen samples (live cells or tissue homogenates) were co-cultured in T-25 flasks with five different human cell lines AV3, RD, 1.1B4, VC3, HEK-293 (European Collection of Authenticated Cell Cultures, Porton Down, UK) that express a wide range of enterovirus receptors. Human cell lines were grown in DME/F12 medium supplemented with penicillin/gentamicin and with 10% heat inactivated FBS (Gibco; Thermo Fisher, Rodano, Italy). Cultured cells were checked monthly for mycoplasma contamination (MycoAlert Plus Mycoplasma kit; Euroclone-Lonza, Pero, Italy). For immunofluorescent detection of enterovirus VP1 antigen, cell cultures were prepared in Millicell EZ 4-well glass slides (Merck, Vimodrone, Italy) as described below. At the third passage, the supernatant of cell cultures was used for RNA extraction and RT-PCR. Extracted RNA was reverse transcribed and enterovirus-specific end-point PCR assays were performed using five different primer sets. Capillary electrophoresis (Agilent 2100 Bioanalyzer, Milano, Italy) was used to detect the precise size of amplicons whose sequences were obtained by the Sanger method. For indirect immunofluorescence, cell monolayers were fixed in PBS containing 4% paraformaldehyde. Enterovirus-infected cells were spotted by staining with two different mouse monoclonal antibodies against the VP1 enteroviral capsid antigen_(9D5 from Merck; 6-E9/2 from Creative Diagnostics). The two antibodies bind to distinct stretches of the VP1 protein. Both recognize acute and persistent enterovirus infection in cultured cells, are devoid of neutralizing activity, and are specific for a vast spectrum of EV types. The 9D5 antibody binds to the consensus motif SIGNAYSMFYDG (37) while 6-E9/2 recognizes the same epitope of the 5D8/1 antibody (38). Alexa Fluor 488-goat anti-mouse IgG was used as secondary antibody.

### Statistical analysis

Statistical analyses were performed using SPSS 25.0 for Windows. Frequency comparisons was performed with the Pearson’s Χ^2^ and Fisher’s exact tests. When comparing donor groups, the significant p values were corrected for multiple comparisons by multiplying the raw p-value by the number of comparisons made (Bonferroni’s correction).

## Results

### Metagenomic sequencing did not detect viruses or other microbes in the pancreas

In the CVB1-spiked quality control samples, the RNA was of high integrity and stability. CVB1 reads were detected with all sequencing approaches used, even at the lowest 10^-8 dilution, confirming high sensitivity of the assays. There was a linear relationship between the spiked-in virus dilution level and the CVB1 reads count (**ESM Fig. 1, ESM Table 2**). In the negative control, we detected one pair of CVB1 reads; this does not meet the species detection criteria and was considered as possible low level cross contamination from the highest viral load samples.

**Table 2.**
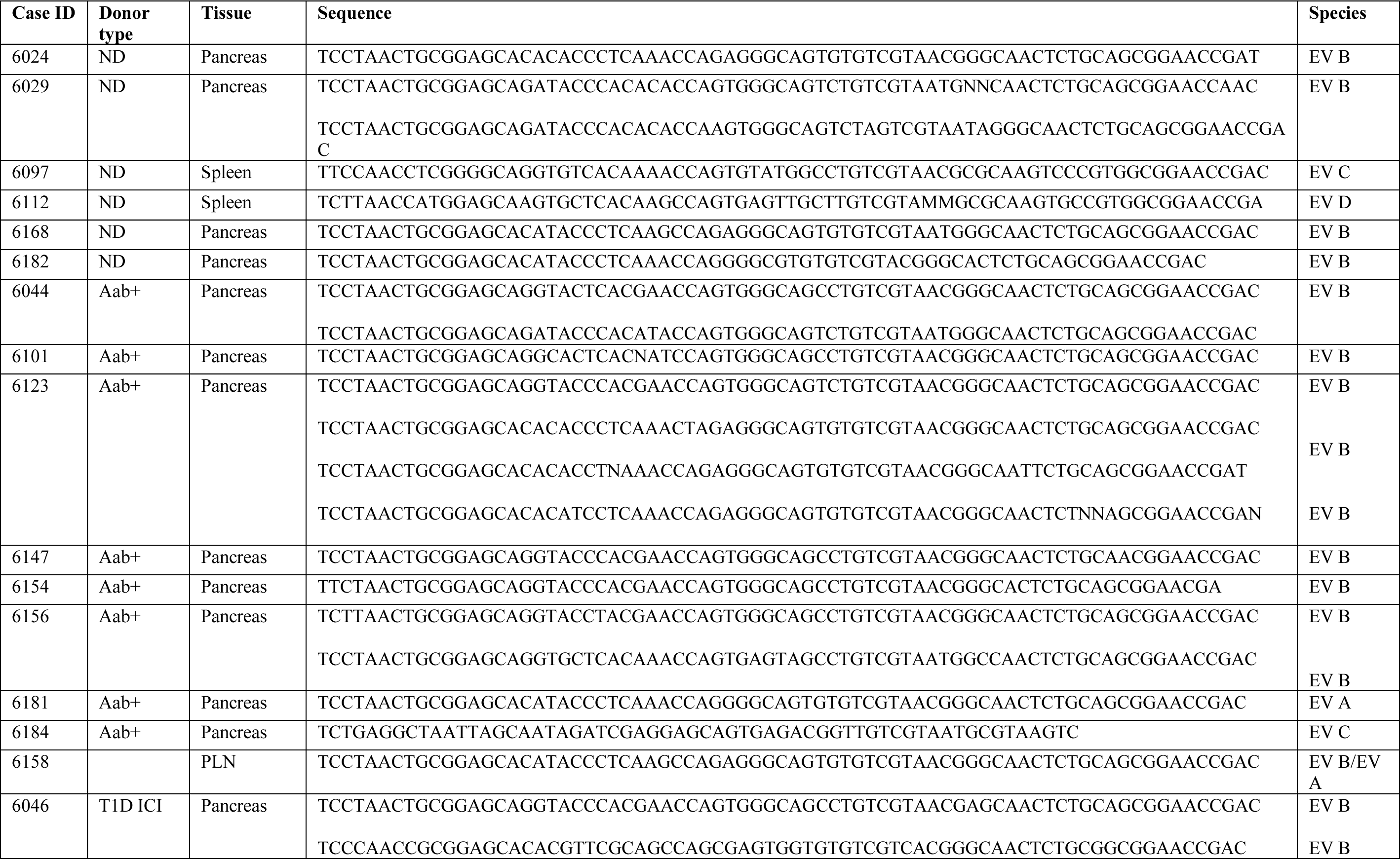
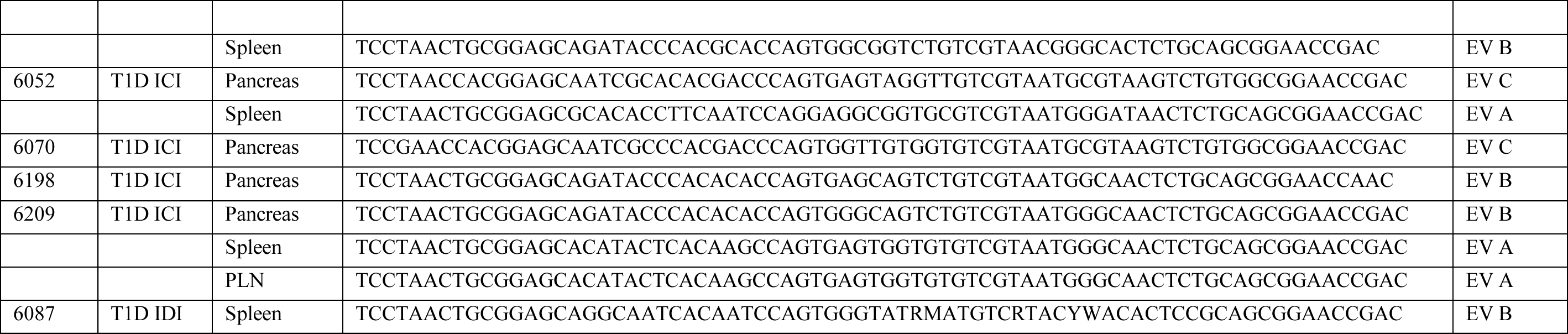
Examples of enterovirus sequences detected in the samples by enterovirus specific RT-PCR and their alignment to different enterovirus species. The amplified region locates within the conserved region of the viral genome and does not allow identification of the exact type of detected enteroviruses. ND, non-diabetic; Aab+, single autoantibody-positive; T1D-ICI, T1D with residual insulin-containing islets; T1D-IDI, T1D with insulin-deficient islets.

Most pancreas samples used at UCL for metagenomic *steps I-IV* sequence analyses had moderately to highly degraded RNA. The search for enteroviral reads did not yield positive hits in any of the analysis steps. In *steps I and II*, the deduplicated reads were mostly human sequences. Taxonomic classification of the “microbial” reads with metaMix resulted in a similar profile for all samples, showing no difference between case and control donors. Most reads were assigned to Enterobacteria phage phiX174, the positive control for Illumina sequencing. The rest of the reads were divided between various environmental bacteria and the “unknown” bin (**ESM Table 3**). In *step III*, which included a specifically designed enterovirus enrichment method, the reads were highly clonal and only a small fraction of the data was non-human. Community profiling identified human and enterobacterial phage phiX sequences, and most of the small number of “microbial” reads were unassigned (representative summary in **ESM Table 4**). In 2/10 samples there were a few reads from Human Herpesvirus 3. We consider likely that this was a contaminant, as this virus is frequently sequenced on the Illumina machines at UCL.

In the RNA extracted from laser microdissected islets (*step IV* analyses with the targeted enterovirus baits), the number of “microbial” reads was low, despite the high sequencing depth. The metaMix profile consisted of 2-3 species per sample and there was low level carry over contamination with viruses routinely sequenced at UCL. To rule out the remote possibility that unassigned reads originate from a species not present in our database, a BLASTn search was performed against the nucleotide-NR database. The reads remained unclassified and only a few thousand reads matched human, bacterial or uncultured eukaryote sequences.

At BCM, 68 pancreatic tissue samples were sequenced, including pooled laser captured islets from three samples, to amplify islet-specific signal in selected donors but no virus sequences were detected.

### Detection of enterovirus RNA using sensitive RT-PCR assays

Analyses of pancreas samples from 137 donors by sensitive RT-PCR methods detected enteroviruses in 3/5 donor groups, and most frequently in the AAb+ donors. A sample was considered positive if at least one of the two laboratories detected enterovirus RNA and the sequence of the PCR product matched with enterovirus. Taken together, enterovirus was detected in the pancreas in 16% (5/32) of T1D-ICI donors, 0% (0/34) of T1D-IDI donors, 0% (0/7) of AAb++ donors, 53% (8/15) of AAb+ donors and 8% (4/49) of ND donors (**Fig. 2 A**). AAb+ donors had significantly higher frequency of positivity compared to T1D-IDI (p-value corrected for multiple comparisons <0.001) and ND donors (corrected p-value 0.004) (see also **ESM Table 5**). Sequencing of the RT-PCR products identified variations in the amplified genome region, implying the presence of different enterovirus genotypes across donors. Based on the obtained sequences over 90% of them belonged to *Enterovirus B* species. However, since the amplified region locates within the conserved part of the viral genome, the identification of the exact genotype of these viruses was not possible. The sequences obtained from all the samples are listed in **Table 2**.

**Figure 2.**
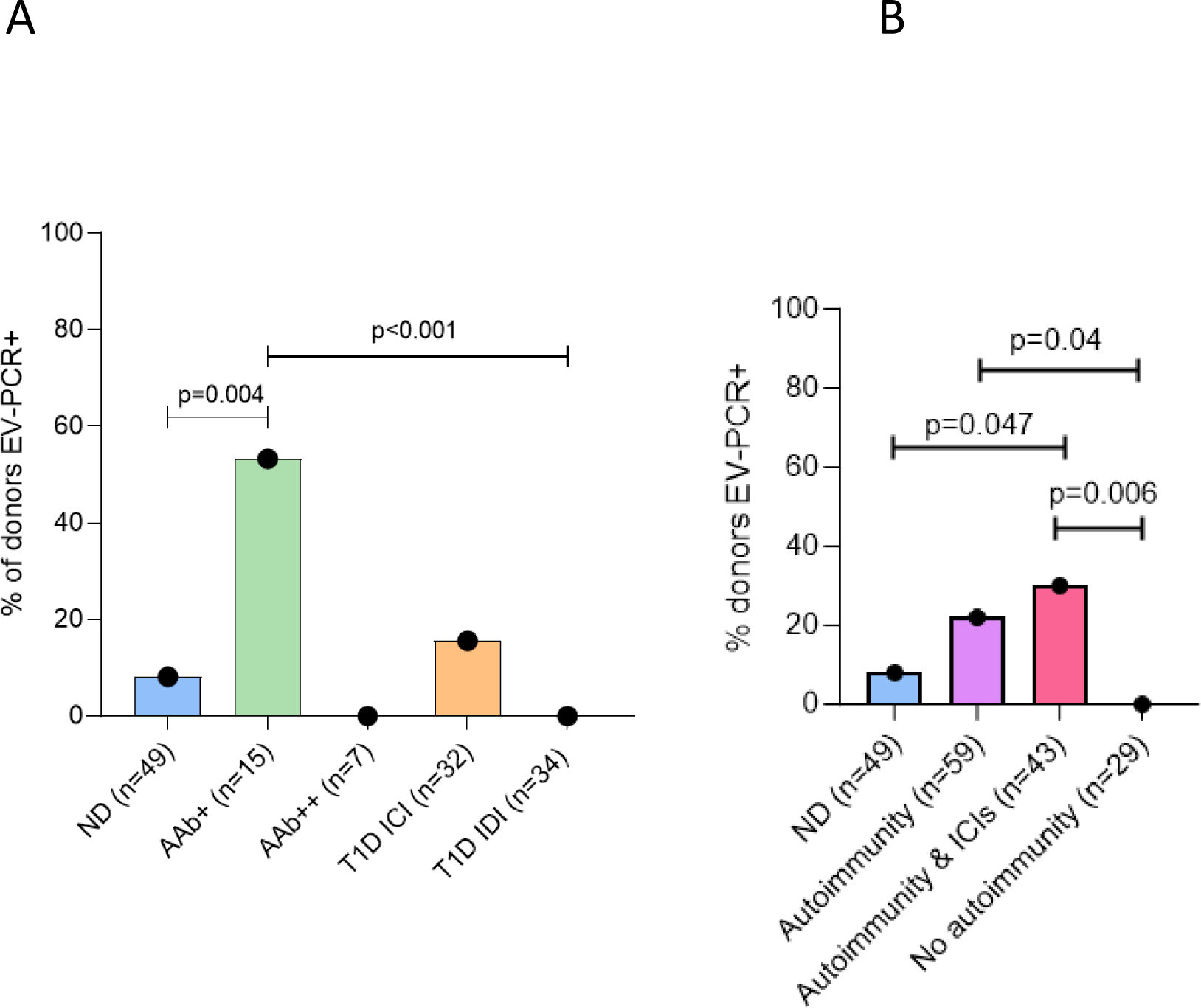
Enterovirus RNA was detected in pancreas with RT-PCR. **A)** Enterovirus RNA was detected in the pancreas across different donor groups. Detection rate was higher in AAb+ donors compared to non-diabetic donors and T1D donors without insulin (T1D-IDI). Significant p-values (corrected for multiple (N=10) comparisons) are shown. Other corrected group comparisons were non-significant. **B)** Donors with active islet autoimmunity (‘Autoimmunity’), as marked by autoantibody-positivity, regardless of T1D status, had higher prevalence of EV RNA positivity, especially those with residual beta cells (‘Autoimmunity & ICIs’), compared to T1D donors without active autoimmunity (‘No autoimmunity’) and non-diabetic control donors (‘ND’). Significant p-values (corrected for multiple (N=6) comparisons).

We then investigated whether the presence of enterovirus RNA in pancreas is associated with active islet autoimmunity, as suggested by the presence of circulating islet autoantibodies in the donors at the time of passing. As a result, 22% (13/59) of the donors with active islet autoimmunity (those without diabetes expressing one or more autoantibodies and those with type 1 diabetes still expressing autoantibodies) carried enterovirus RNA compared to 0% (0/29) of donors with type 1 diabetes lacking autoantibodies (corrected p-value 0.04) and 8% (4/49) of autoantibody negative non-diabetic donors (p-value not significant). In addition, when taking only those donors with active islet autoimmunity and residual beta cells (including T1D with ICI, AAb+ and AAb++ donors), 30% (13/43) were enterovirus positive (vs. non-diabetic donors, corrected p-value 0.047; vs. T1D without islet autoimmunity, corrected p-value 0.006) (**Fig. 2, B, ESM Table 6**).

Paired pancreas and pancreatic lymph node samples from seven donors were available to examine enterovirus RNA using RT-PCR. One T1D-ICI donor tested positive in the pancreas and two donors (1 T1D-ICI and 1 AAb++) in the PLN (the T1D-ICI donor was also positive in the pancreas). Five donors tested negative in both tissues (2 T1D-ICI, 2 AAb++ and 1 T1D-IDI).

In spleen, enterovirus RNA was detected in 9% (9/97) of the donors (4/36 of ND; 0/12 of AAb+; 1/7 of AAB++; 3/19 of T1D-ICI; 1/23 of T1D-IDI) showing no statistically significant differences between groups (**Fig. 3A, ESM Table 7**). We did not detect enterovirus RNA in duodenal tissue of the 65 donors tested.

**Figure 3.**
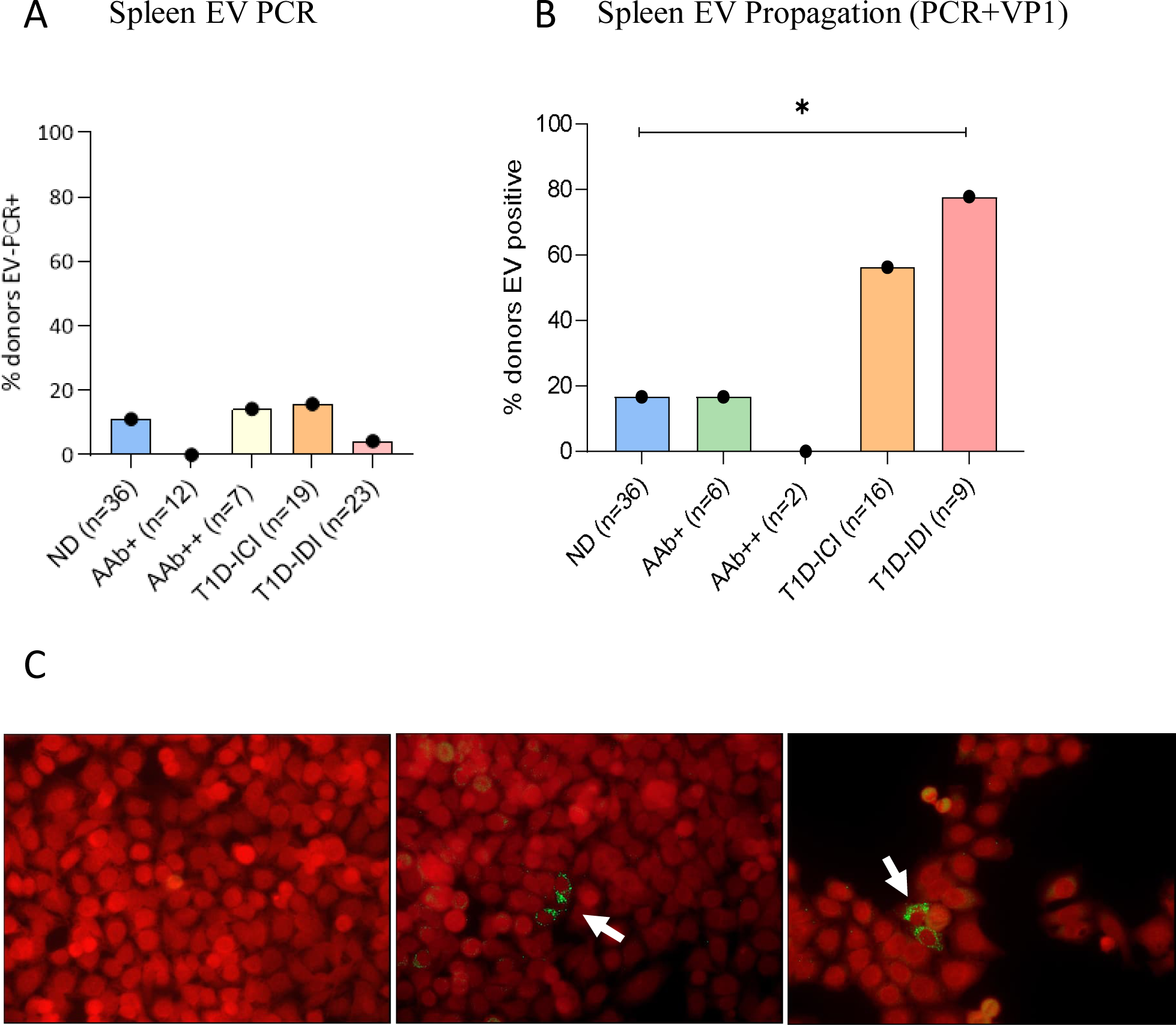
Enterovirus detection in spleen. A) Enterovirus RNA followed by sequencing was detected in 9/97 samples using RT-PCR, without statistical significance between the donor groups. B) Enterovirus propagation in cultured cells: combined results of the RT-PCR and immunostaining methods yielded 23/69 of the samples positive for enterovirus, with type 1 diabetic donors without residual insulin-containing islets (T1D-IDI) being significantly more positive compared to non-diabetic (ND) donors (p=0.01). Fisher exact test 2-sided. Significant p-values corrected for multiple (N=10) comparisons. C) Immunostaining of cultured AV3 cells inoculated with spleen homogenates to visualize the production of enterovirus VP1 capsid protein (mouse anti-enterovirus monoclonal antibody clone 6-E9/2, Creative Diagnostics). Left panel: cell culture inoculated by a spleen homogenate from a non-diabetic donor: production of viral protein is not observed. Middle and right panels: results obtained by spleen homogenates from two donors with type 1 diabetes showing the production of enterovirus protein in a few cultured cells (indicated by green color and white arrows). Virus replication did not lead to a cytopathic effect typically observed in acute enterovirus infections. Magnification 250x.

The quality of extracted RNA was studied in selected samples. The RNA Quality Number (RQN) values varied considerably between donors ranging from 1 (totally degraded RNA) to 10 (intact RNA) and tended to be higher in pancreas than in spleen and duodenum. The median RQN values were 4.5 (range 1-7.8) in pancreas (N=12), 1.6 (1-6.2) in spleen (N=5) and 2.0 (1-7.3) in duodenum (N=10) samples. A low RQN score did not seem to affect the enterovirus PCR positivity since positive results were obtained also from samples with poor RNA quality (**ESM Table 8**). In all enterovirus-positive samples, virus loads were very low, often close to the detection limit of the assay.

### Detection of enterovirus in spleen using virus enrichment in human cell lines before RT-PCR

Sixty-nine spleen samples were analysed following virus amplification in cell culture prior to RT-PCR (n=69) and immunostaining steps (n=66). Criterium for positivity was an enterovirus positive result for either one of the methods: a) for RT-PCR meaning Ct-value below 32 plus amplicons of the expected size by capillary electrophoresis, and/or b) cytoplasmic fluorescent staining in cultured cells. RT-PCR found positivity in 23/69 samples and immunostaining in 19/66 samples analysed. The agreement between methods was 97%, both being positive in 19 and negative in 46 of 66 samples analysed. Two samples were positive only by RT-PCR. In such cases the result was deemed positive. Overall, enteroviruses were detected in 33% (23/69) of spleen samples. This is a significantly higher rate than that obtained by direct RT-PCR analysis of frozen spleen samples without virus propagation in cultured cells (above). In total, 56% (9/16) of T1D-ICI donors, 78% (7/9) of T1D-IDI donors, 0% (0/2) of AAb++ donors, 17% (1/6) of AAb+ donors and 17% (6/36) of ND donors, were enterovirus positive using this protocol (**Fig. 3B, ESM Table 9**). T1D IDI donors had significantly higher frequency of positivity compared to ND donors (p=0.01). T1D ICI donors trended to be more frequently enterovirus-positive than ND donors, without reaching statistical significance (P=0.069). When combining the direct RT-PCR from spleen with the detection of enterovirus replication in cell culture (replication detected by either RT-PCR or immunostaining, or both) (134 donors), there was a trend towards donors with type 1 diabetes being more positive for enteroviruses compared to other donor groups. Enterovirus RNA was found in 39% (10/26) of T1D-ICI donors, 30% (8/27) of T1D-IDI donors, 14% (1/7) of AAb++ donors, 8% (1/12) of AAb+ donors and 13% (8/62) of ND donors. Importantly, enterovirus propagation from spleen cells yielded viruses which did not cause an evident cytopathic effect in cultured cells, unlike what is typically observed with replication competent viruses that cause acute enterovirus infections. In cell cultures, only a few cells stained positive for enterovirus capsid protein VP1 indicating limited replication of the virus (**Fig. 3C**).

## Discussion

In the coordinated effort of the nPOD-Virus Group, we conducted the largest screening for RNA viruses and specifically enterovirus in the pancreas of organ donors with evidence of islet autoimmunity and/or type 1 diabetes. We employed multiple approaches to detect viral RNA. Studies aiming at detecting viral proteins are reported in companion manuscripts. For the first time in this setting, we employed two metagenomic sequencing methods to broadly ascertain the possible presence of RNA viruses, and based on previous literature associations, we employed enterovirus specific methods (RT-PCR followed by sequencing of PCR amplicons to detect enterovirus RNA, and *in vitro* virus propagation in cell culture followed by RT-PCR plus immunostaining to detect enterovirus replication).

We developed two non-biased RNA-Seq methods specifically to analyse organ donor pancreas, which has never been attempted before. Despite all the measures we took to maximize sensitivity and that our sequencing approaches had relatively high sensitivity in detecting enterovirus RNA in infected cell lines, both RNA-Seq methods, even when using sequence capture or RNA from laser-captured islets, failed to detect any viral RNA sequences. While we examined transplant grade pancreata, we consider that several factors could have negatively impacted assay sensitivity to detect extremely low amounts of viral RNA. These include: 1) signal loss due to RNA degradation in organ donor pancreas, an organ that is known to degrade rapidly; 2) the well-known rarity of infected cells with associated sampling limitations, and thus likely suboptimal targeting of sampling; 3) the limited likelihood that we may detect evidence of viral infection at the time of passing, since such an infection may have occurred years prior to disease development; 4) the possible presence of replication-defective viruses producing too small amounts of RNA copies (14,39–41). This experience provides a backbone for improvement for future studies likewise aiming at detecting viruses in pancreas.

In contrast, our highly sensitive RT-PCR assays detected enterovirus RNA in the pancreas across all donor groups. Even though the amount of virus was very low and close to the detection limit, two independent laboratories confirmed the presence of enterovirus RNA in the pancreas of 17 donors, overall. Enterovirus RNA was more prevalent in non-diabetic donors with a single autoantibody compared to control donors, which may represent persons at earlier stages of the disease progression, when enteroviruses are suggested to act as triggers and may be more likely to be present. Moreover, donors with signs of active autoimmunity, including islet autoantibody positive with or without type 1 diabetes, had higher prevalence of enterovirus RNA positivity than donors without active autoimmunity. Thus, it is possible that enterovirus infection in the pancreas is associated with ongoing autoimmune responses against beta cell antigens. The fact that the RNA virus load was extremely low fits with a low-grade, possibly persistent, infection rather that an acute infection. Beta-cells may be particularly permissive for such prolonged/persistent infection due their relatively weak anti-viral innate immune response (13) and high expression of CAR (11) used by CVB group enteroviruses.

However, in the comparison of the different donor groups we acknowledge several limitations: 1) the limited number of enterovirus RNA positive donors; 2) the varied interval time between autoantibody conversions or diagnosis and passing; 3) sampling limitations since the disease process occurs without uniformity across the pancreas; 4) the extremely low amounts of virus which may also have caused stochastic variation in the amplification of viral RNA using RT-PCR. Additional variation could be caused by pancreatic enzymes and other compounds that degrade RNA, particularly when the viral RNA is present in unencapsidated forms where it is not protected by capsid proteins. This may make it difficult to detect a persisting enterovirus infection in pancreas (41), since such viruses replicate slowly and generate much reduced amounts of complete virions compared to the acute infection (42). When evaluating the RNA quality in a subset of pancreas, spleen and duodenal samples, variation in RNA degradation levels was evident. However, poor RQN values didn’t seem to affect the detection of enterovirus sequences by RT-PCR. This is not surprising, since the employed RT-PCR methods amplify very short fragments of viral RNA (in the range of 90 to 200bp). The impact of RNA degradation is expected to be greater for RNA-Seq methods.

This study shows, for the first time, that enteroviruses can be found in the PLNs as well. In fact, this fits well with their draining function as the lymph flow from the infected pancreas can carry virus to the nodes. Enterovirus RNA was also found in the spleen, another important lymphoid organ. Previous studies have shown elevated enterovirus titres in spleen during acute infection in human beings and in animals (43–45), and in mouse models the spleen is enterovirus-positive during a later phases of infection (40,42,46). Detection of enterovirus RNA in spleen suggests infection of lymphoid cells. This is also supported by our own finding showing that enterovirus RNA can be detected using *in situ* RNA probes in immune cells infiltrating islets in the pancreas with type 1 diabetes (47). We conducted co-culture studies in which spleen-derived enteroviruses caused no cytopathic effects in cell culture; this behaviour was previously seen with persistent enterovirus strains and linked to replication-defective viruses, including studies showing that CVB3 can lead to persistent and replication-defective infection in the mouse pancreas, accompanied by deletion in the 5’-non-coding region of the viral genome which regulate viral replication (14,48).

Progression of the infection in our co-culture studies was slow and less than 5% cells stained positive for the enterovirus VP1 protein (Fig. 3C), further supporting the presence of strains that produce persisting infection and/or replication-defective viruses in the spleen. The detection of virus in the lymphoid tissues of donors with type 1 diabetes raises new questions: it will be important to discover which cell types are enterovirus-positive in spleen (presumably of lymphoid nature) and how the virus may alter the function of infected cells. Previous studies have shown that enterovirus can infect human and murine leukocytes (49–51). In our recent study of patients with type 1 diabetes and controls, enterovirus RNA was found significantly more often in PBMC subsets than in plasma and virus detection correlated with islet autoimmunity and the IFIH1 genotype (52).

Enterovirus RNA was not detected in duodenal samples, in contrast with a previous study where both enterovirus RNA and VP1 protein were detected in duodenal biopsies taken from living patients with type 1 diabetes or near diabetes diagnosis (16). In addition, enterovirus VP1 protein was detected in formalin-fixed paraffin-embedded duodenal samples collected from these same nPOD donors as described in the accompanying publication (Rodriguez-Calvo et al). On the other hand, a previous study failed to find enterovirus RNA in duodenal biopsies taken from patients with type 1 diabetes but detected enterovirus protein at low frequency (18). The reason of these discrepancies is unknown. Methodological differences, variation in the selection of cases, sample preparation, and limited sampling may be involved. For example, previous studies have been based on biopsy samples collected from living patients while the present study examined tissues from cadaver organ donors. Unlike pancreas tissue from organ donors, duodenal specimens are not perfused with tissue preservation buffer after harvesting, to protect from cold ischemia. Thus, unfixed duodenal tissue may not have been optimally preserved during the transport, leading to partial RNA degradation.

A limitation of our study is that the RT-PCR approach did not allow the identification of the infected cell types in pancreas or other organs. Thus, the cell types harbouring enterovirus RNA remain unknown. A previous study that used *in situ* hybridization in tissue sections showed enterovirus RNA in pancreatic islets of some donors with type 1 diabetes (4). Another study showed enterovirus RNA by RT-PCR in cultured islets that had been isolated from living type 1 diabetes patients (6). A recent study of nPOD donors found enterovirus RNA in both insulin positive and negative cells in some donors with type 1 diabetes, including immune cells in the pancreas using fluorescent *in situ* hybridization (47). It is well known and further corroborated by the accompanying nPOD-Virus group paper (Rodriguez-Calvo et al) that enterovirus-VP1 protein is found primarily in insulin-positive beta cells and in the spleen in immune cells. These cells are quite rare and this may explain the technical difficulties in detecting enterovirus RNA in pancreas (1). An additional limitation is that the exact virus genotypes could not be determined since RT-PCR amplified a highly conserved genome region. However, we propose that even partial identification is valuable as it further links enterovirus infections to the pancreas and type 1 diabetes. It should also be noted that some control subjects who did not have type 1 diabetes and who were negative for islet autoantibodies were positive for enterovirus RNA in the pancreas, spleen or lymph nodes. This is not surprising given the high prevalence of enterovirus infections in the general population (53). Additional data from our group suggest that host responses may be critical, among other factors (genetics, virus variants) in modulating the outcome of low-grade enterovirus infections in the pancreas.

Together with other studies of the nPOD-Virus Group, the present findings demonstrate that enterovirus RNA is present in organ donors with islet autoimmunity and insulin containing islets, whether at the preclinical stage or after diagnosis, with an increased frequency compared to donors without diabetes. Moreover, donors with a single autoantibody had the highest prevalence of detection, which would be consistent with enterovirus infections occurring early in the natural history of the disease. Despite limitations, the data support an association of enterovirus RNA with islet autoimmunity and suggest a low-grade enteroviral infection of pancreatic and lymphoid tissues.

## Acknowledgements

The authors wish to thank all families associated with organ donation for research purposes for their gift that made efforts like those in this program possible. In addition, appreciation is given to the OPO who make the nPOD program possible.

The authors would like to thank J.Ilomäki, T.Kuusela, and M.Ovaskainen, Faculty of Medicine and Health Technology, Tampere University, Tampere, Finland, for technical assistance.

## Data availability

Data generated and analysed during this study are available through the corresponding author upon request.

## Contribution statement

AP, SO, JEL, HH, VP, JFP and AT were responsible for the overall design of the study. SO was responsible for the virus PCR and sequencing analyses at Tampere laboratory, and AR and REL for the virus PCR and sequencing analyses at BCM. SM, DD, ICG, JB and VP contributed to the RNA-Seq analyses at UCL. MCR and JFP, were responsible for the whole genome shotgun sequencing at BCM. AT was responsible for the virus propagation studies at the Varese laboratory. MO and JEL performed the overall data analyses. JEL, SO, SM and MO were the primary contributors in the preparation of the manuscript. All authors contributed to data interpretation and discussion, reviewed the manuscript and approved the final version. JEL is the guarantor of this work and, as such, has full access to the data in the study and takes responsibility for the integrity of the data and the accuracy of the data analysis.

## Abbreviations

Aab: autoantibody
CAR: coxsackie-adenovirus receptor
CVB: coxsackievirus B
DiViD: Diabetes Virus Detection study
ds: double-stranded
ICI: insulin-containing islet
IDI: insulin-deficient islet
ND: non-diabetic
nPOD: network for Pancreatic Organ donors with Diabetes
VP1: viral capsid protein 1
WGS: whole genome shotgun

