## Supplemental material for "Detection of enterovirus RNA in pancreas and lymphoid tissues of organ donors with type 1 diabetes"

Electronic supplementary material

Laiho JE et al

**ESM Table 1: Donor demongraphics including RRiD, age, sex, BMI, c-peptide, duration of disease, AAb status, Ethnicity and which tissue types were studied.**

| Study Number | RRiD | Donor Type | Age (years) | Sex | BMI (kg/m2) | C-peptide (ng/ml) | Disease Duration | Autoantibodies (Aabs) | Ethnicity | EV-PCR Pancreas | EV-PCR Spleen | EV-PCR Duodenum | EV-PCR PLN | RNA-Seq Pancreas | EV-Propagation Spleen |
|---|---|---|---|---|---|---|---|---|---|---|---|---|---|---|---|
| 6005 | SAMN15879062 | ND | 0-10 | Female | 15.7 | | NA | NA | Caucasian | | Y | | | | |
| 6009 | SAMN15879066 | ND | 40-50 | Male | 30.6 | 11.32 | NA | NA | Caucasian | Y | | | | | |
| 6010 | SAMN15879067 | ND | 40-50 | Female | 19.7 | | NA | NA | Caucasian | Y | Y | | | | |
| 6012 | SAMN15879069 | ND | 60-70 | Female | 23.7 | 2.97 | NA | NA | Caucasian | Y | | | | Y | |
| 6013 | SAMN15879070 | ND | 60-70 | Male | 24.2 | 2.8 | NA | NA | Caucasian | Y | | | | | |
| 6016 | SAMN15879073 | ND | 60-70 | Female | 31.2 | | NA | NA | Caucasian | Y | | | | | |
| 6017 | SAMN15879074 | ND | 50-60 | Female | 24.8 | 9.89 | NA | NA | Caucasian | Y | | | | Y | |
| 6019 | SAMN15879076 | ND | 40-50 | Male | 31 | 0.47 | NA | NA | Caucasian | Y | | | | Y | |
| 6020 | SAMN15879077 | ND | 60-70 | Male | 29.8 | 2.82 | NA | NA | Caucasian | Y | | | | | |
| 6022 | SAMN15879079 | ND | 70-80 | Male | 30.6 | 4.99 | NA | NA | Caucasian | Y | | | | | |
| 6024 | SAMN15879081 | ND | 20-30 | Male | 27.8 | 3.52 | NA | NA | Caucasian | Y | Y | | | Y | |
| 6029 | SAMN15879086 | ND | 20-30 | Female | 22.6 | | NA | NA | Hispanic | Y | Y | | | | |
| 6030 | SAMN15879087 | ND | 30-40 | Male | 27.1 | 2.54 | NA | NA | Caucasian | Y | Y | | | Y | |
| 6034 | SAMN15879091 | ND | 30-40 | Female | 25.2 | 3.15 | NA | NA | Caucasian | Y | Y | | | Y | |
| 6047 | SAMN15879104 | ND | 0-10 | Male | 23.9 | 0.65 | NA | NA | Caucasian | Y | | | | Y | Y |
| 6073 | SAMN15879130 | ND | 10-20 | Male | 36 | 0.69 | NA | NA | Caucasian | Y | | | | Y | |
| 6075 | SAMN15879132 | ND | 10-20 | Male | 14.9 | 2.94 | NA | NA | African Am | Y | | | | Y | |
| 6095 | SAMN15879152 | ND | 30-40 | Male | 35.5 | | NA | NA | Hispanic | Y | Y | Y | | Y | |
| 6096 | SAMN15879153 | ND | 10-20 | Female | 18.8 | 2.97 | NA | NA | African Am | Y | Y | Y | | Y | |
| 6097 | SAMN15879154 | ND | 40-50 | Female | 36.4 | 16.76 | NA | NA | Caucasian | Y | Y | Y | | | |
| 6098 | SAMN15879155 | ND | 10-20 | Male | 22.8 | 1.41 | NA | NA | Caucasian | Y | | | | Y | |
| 6099 | SAMN15879156 | ND | 10-20 | Male | 30 | 5.37 | NA | NA | Caucasian | Y | | | | Y | |
| 6102 | SAMN15879159 | ND | 40-50 | Female | 35.1 | 0.55 | NA | NA | Caucasian | Y | Y | Y | | Y | Y |
| 6103 | SAMN15879160 | ND | 0-10 | Male | 16.8 | 0.98 | NA | NA | Caucasian | Y | Y | Y | | Y | |

| | | | | | | | | | | | | | | | |
|---|---|---|---|---|---|---|---|---|---|---|---|---|---|---|---|
| 6104 | SAMN15879161 | ND | 40-50 | Male | 20.5 | 20.55 | NA | NA | Caucasian | Y | Y | Y | | Y | Y |
| 6106 | SAMN15879163 | ND | 0-10 | Male | 17.4 | 7.36 | NA | NA | Caucasian | Y | Y | Y | | | |
| 6112 | SAMN15879169 | ND | 0-10 | Female | 18.4 | 5.11 | NA | NA | Hispanic | Y | Y | Y | | | |
| 6117 | SAMN15879174 | ND | 0-10 | Male | 18.4 | 3.27 | NA | NA | Caucasian | | | | | | Y |
| 6126 | SAMN15879183 | ND | 20-30 | Male | 25.1 | 0.88 | NA | NA | Hispanic | Y | Y | Y | | Y | |
| 6130 | SAMN15879187 | ND | 0-10 | Male | 18.5 | 4.8 | NA | NA | Caucasian | Y | Y | Y | | | |
| 6131 | SAMN15879188 | ND | 20-30 | Male | 24.8 | 1.01 | NA | NA | Caucasian | Y | Y | Y | | | |
| 6137 | SAMN15879194 | ND | 0-10 | Female | 24.2 | 12.13 | NA | NA | Hispanic | Y | Y | Y | | Y | Y |
| 6140 | SAMN15879197 | ND | 30-40 | Male | 21.7 | 11.1 | NA | NA | Caucasian | Y | Y | Y | | | Y |
| 6160 | SAMN15879216 | ND | 20-30 | Male | 23.9 | 0.4 | NA | NA | Caucasian | Y | Y | Y | | Y | Y |
| 6162 | SAMN15879218 | ND | 20-30 | Male | 28.9 | 7.61 | NA | NA | African Am | Y | Y | Y | | | |
| 6165 | SAMN15879221 | ND | 40-50 | Female | 25 | 4.45 | NA | NA | Caucasian | Y | Y | Y | | Y | |
| 6168 | SAMN15879224 | ND | 50-60 | Male | 25.2 | | NA | NA | Hispanic | Y | Y | Y | | | |
| 6172 | SAMN15879228 | ND | 10-20 | Female | 32.4 | 8.02 | NA | NA | Caucasian | Y | Y | Y | | | |
| 6174 | SAMN15879230 | ND | 20-30 | Male | 19.5 | 3 | NA | NA | Caucasian | Y | Y | Y | | | |
| 6178 | SAMN15879234 | ND | 20-30 | Female | 27.5 | 4.55 | NA | NA | Caucasian | Y | | | | | Y |
| 6179 | SAMN15879235 | ND | 20-30 | Female | 20.7 | 2.74 | NA | NA | Caucasian | Y | Y | Y | | | |
| 6182 | SAMN15879238 | ND | 0-10 | Male | 26 | 2.28 | NA | NA | Caucasian | Y | Y | Y | | Y | Y |
| 6190 | SAMN15879246 | ND | 0-10 | Male | 14.2 | 5.5 | NA | NA | Hispanic | | | | | | Y |
| 6227 | SAMN15879283 | ND | 10-20 | Female | 26.4 | 2.75 | NA | NA | Caucasian | | | | | | Y |
| 6238 | SAMN15879294 | ND | 20-30 | Male | 21.7 | 1.17 | NA | NA | African Am | Y | Y | | | | |
| 6254 | SAMN15879310 | ND | 30-40 | Male | 30.5 | 6.43 | NA | NA | Caucasian | Y | Y | | | | Y |
| 6278 | SAMN15879332 | ND | 10-20 | Female | 21.3 | 4.54 | NA | NA | African Am | Y | Y | | | | |
| 6282 | SAMN15879336 | ND | 10-20 | Male | 41.9 | 6.83 | NA | NA | Caucasian | | | | | | Y |
| 6289 | SAMN15879343 | ND | 10-20 | Male | 38.3 | 8.05 | NA | NA | African Am | | | | | | Y |
| 6295 | SAMN15879349 | ND | 40-50 | Female | 30.4 | 10.91 | NA | NA | African Am | | | | | | Y |
| 6318 | SAMN15879372 | ND | 0-10 | Female | 17.6 | 3.89 | NA | NA | Caucasian | | | | | | Y |
| 6333 | SAMN15879387 | ND | 20-30 | Female | 24.9 | 9.37 | NA | NA | Caucasian | | | | | | Y |
| 6338 | SAMN15879392 | ND | 10-20 | Male | 22.8 | 5.01 | NA | NA | African Am | | | | | | Y |
| 6339 | SAMN15879393 | ND | 20-30 | Male | 25 | 10.56 | NA | NA | Caucasian | Y | Y | | | | |

| | | | | | | | | | | | | | | | |
|---|---|---|---|---|---|---|---|---|---|---|---|---|---|---|---|
| 6340 | SAMN15879394 | ND | 0-10 | Male | 20.3 | 3.88 | NA | NA | Caucasian | | | | | | Y |
| 6350 | SAMN15879404 | ND | 0-10 | Female | 12.7 | 2.21 | NA | NA | Hispanic | | | | | | Y |
| 6353 | SAMN15879406 | ND | 10-20 | Male | 28.3 | 1.76 | NA | NA | African Am | | | | | | Y |
| 6356 | SAMN15879409 | ND | 0-10 | Female | 17.1 | 1.65 | NA | NA | Caucasian | | | | | | Y |
| 6357 | SAMN15879410 | ND | 0-10 | Male | 15.3 | 8.82 | NA | NA | Caucasian | | | | | | Y |
| 6364 | SAMN15879417 | ND | 0-10 | Male | 18 | 3.3 | NA | NA | Hispanic | | | | | | Y |
| 6366 | SAMN15879419 | ND | 20-30 | Female | 20.5 | 0.41 | NA | NA | Hispanic | | | | | | Y |
| 6368 | SAMN15879421 | ND | 30-40 | Male | 20.7 | 3.05 | NA | NA | Caucasian | | | | | | Y |
| 6369 | SAMN15879422 | ND | 40-50 | Male | 18.8 | 6.42 | NA | NA | Caucasian | | | | | | Y |
| 6375 | SAMN15879428 | ND | 20-30 | Male | 31.8 | 17.32 | NA | NA | Caucasian | Y | Y | | | | Y |
| 6381 | SAMN15879434 | ND | 0-10 | Male | 22.6 | 5.6 | NA | NA | African Am | | | | | | Y |
| 6384 | SAMN15879437 | ND | 10-20 | Male | 18.2 | 0.7 | NA | NA | Caucasian | Y | Y | | | | |
| 6385 | SAMN15879438 | ND | 10-20 | Male | 16.2 | 1.55 | NA | NA | Caucasian | | | | | | Y |
| 6386 | SAMN15879439 | ND | 10-20 | Male | 23.9 | 1.12 | NA | NA | Caucasian | | | | | | Y |
| 6401 | SAMN15879454 | ND | 20-30 | Female | 31.3 | 12.81 | NA | NA | Hispanic | Y | Y | | | | |
| 6406 | SAMN15879459 | ND | 0-10 | Male | 16.8 | 4.07 | NA | NA | Caucasian | Y | Y | | | | Y |
| 6407 | SAMN15879460 | ND | 0-10 | Female | 16 | 5.35 | NA | NA | Caucasian | | | | | | Y |
| 6412 | SAMN15879465 | ND | 10-20 | Female | 24 | 45.57 | NA | NA | Caucasian | | | | | | Y |
| 6413 | SAMN15879466 | ND | 10-20 | Female | 19 | 5.27 | NA | NA | Caucasian | Y | Y | | | | Y |
| 6420 | SAMN15879473 | ND | 10-20 | Male | 15.4 | 1.27 | NA | NA | Caucasian | | | | | | Y |
| 6027 | SAMN15879084 | AAb+ | 10-20 | Male | 19.9 | | NA | ZnT8A+ | Caucasian | Y | | | | Y | |
| 6044 | SAMN15879101 | AAb+ | 40-50 | Male | 27.4 | 13.55 | NA | GADA+ | Hispanic | Y | Y | | | Y | |
| 6090 | SAMN15879147 | AAb+ | 0-10 | Male | 18.8 | 5.34 | NA | GADA+ | Hispanic | Y | Y | Y | | Y | |
| 6101 | SAMN15879158 | AAb+ | 60-70 | Male | 34.3 | 26.18 | NA | GADA+ | Caucasian | Y | Y | Y | | Y | |
| 6123 | SAMN15879180 | AAb+ | 20-30 | Female | 17.6 | 2.01 | NA | GADA+ | Caucasian | Y | Y | Y | | Y | Y |
| 6147 | SAMN15879203 | AAb+ | 20-30 | Female | 32.9 | 3.19 | NA | GADA+ | Caucasian | Y | | | | Y | |
| 6151 | SAMN15879207 | AAb+ | 30-40 | Male | 24.2 | 5.49 | NA | GADA+ | Caucasian | Y | Y | Y | | Y | Y |
| 6154 | SAMN15879210 | AAb+ | 40-50 | Female | 24.5 | 0.049 | NA | GADA+ | Caucasian | Y | Y | Y | | Y | |
| 6156 | SAMN15879212 | AAb+ | 40-50 | Male | 19.8 | 13.34 | NA | GADA+ | Caucasian | Y | Y | Y | | Y | Y |
| 6171 | SAMN15879227 | AAb+ | 0-10 | Female | 14.8 | 8.95 | NA | GADA+ | Caucasian | Y | Y | | | Y | Y |

| | | | | | | | | | | | | | | | |
|---|---|---|---|---|---|---|---|---|---|---|---|---|---|---|---|
| 6181 | SAMN15879237 | AAb+ | 30-40 | Male | 21.9 | 0.06 | NA | GADA+ | Caucasian | Y | Y | Y | | Y | Y |
| 6184 | SAMN15879240 | AAb+ | 40-50 | Female | 27 | 3.42 | NA | GADA+ | Hispanic | Y | Y | Y | | Y | |
| 6314 | SAMN15879368 | AAb+ | 20-30 | Male | 23.8 | 1.49 | NA | GADA+ | Caucasian | Y | | | | | |
| 6400 | SAMN15879453 | AAb+ | 20-30 | Male | 22.2 | 4.17 | NA | GADA+ | Hispanic | Y | Y | | | | |
| 6421 | SAMN15879474 | AAb+ | 10-20 | Male | 17.9 | 1.84 | NA | GADA+ | Hispanic | Y | Y | | | | Y |
| 6080 | SAMN15879137 | AAb++ | 60-70 | Female | 21.3 | 1.84 | NA | mIAA+ GADA+ | Caucasian | Y | Y | Y | Y | Y | Y |
| 6158 | SAMN15879214 | AAb++ | 40-50 | Male | 29.7 | 0.51 | NA | mIAA+ GADA+ | Caucasian | Y | Y | Y | Y | Y | Y |
| 6167 | SAMN15879223 | AAb++ | 30-40 | Male | 26.3 | 5.43 | NA | IA2A+ ZnT8A+ | Caucasian | Y | Y | Y | Y | Y | |
| 6197 | SAMN15879253 | AAb++ | 20-30 | Male | 28.2 | 17.48 | NA | GADA+ IA2A+ | African Am | Y | Y | Y | | Y | |
| 6267 | SAMN15879321 | AAb++ | 20-30 | Female | 23.5 | 16.59 | NA | GADA+ IA2A+ | Caucasian | Y | Y | Y | | | |
| 6424 | SAMN15879477 | AAb++ | 10-20 | Male | 51.4 | 6.97 | NA | mIAA+ GADA+ | Hispanic | Y | Y | | | | |
| 6429 | SAMN15879482 | AAb++ | 20-30 | Male | 19.6 | 2.25 | NA | mIAA+ GADA+ | African Am | Y | Y | | | | |
| 6038 | SAMN15879095 | T1D ICI | 30-40 | Female | 30.9 | 0.2 | 20 | Negative | Caucasian | | Y | | | | |
| 6046 | SAMN15879103 | T1D ICI | 10-20 | Female | 25.2 | nd | 8 | GADA+ ZnT8A+ | Caucasian | Y | Y | | | Y | Y |
| 6049 | SAMN15879106 | T1D ICI | 10-20 | Female | 20.8 | nd | 10 | GADA+ mIAA+ | African Am | Y | | | | | |
| 6051 | SAMN15879108 | T1D ICI | 20-30 | Male | 21.5 | nd | 13 | mIAA+ | Caucasian | Y | | | | Y | |
| 6052 | SAMN15879109 | T1D ICI | 10-20 | Male | 20.3 | 0.18 | 1 | GADA+ mIAA+ | African Am | Y | Y | | | Y | Y |
| 6070 | SAMN15879127 | T1D ICI | 20-30 | Female | 21.6 | nd | 7 | GADA+ mIAA+ | Caucasian | Y | Y | Y | | Y | |
| 6084 | SAMN15879141 | T1D ICI | 10-20 | Male | 26.3 | nd | 4 | mIAA+ | Caucasian | Y | Y | | | Y | Y |
| 6088 | SAMN15879145 | T1D ICI | 30-40 | Male | 27 | nd | 5 | GADA+ IA2A+ mIAA+ ZnT8A+ | Caucasian | Y | Y | Y | | Y | |
| 6113 | SAMN15879170 | T1D ICI | 10-20 | Female | 24.75 | nd | 1.58 | mIAA+ | Caucasian | Y | Y | Y | | Y | Y |
| 6180 | SAMN15879236 | T1D ICI | 20-30 | Male | 25.9 | nd | 11 | GADA+ IA2A+ mIAA+ ZnT8A+ | Caucasian | Y | | | | Y | Y |
| 6195 | SAMN15879251 | T1D ICI | 10-20 | Male | 23.7 | nd | 5 | GADA+ IA2A+ mIAA+ ZnT8A+ | Caucasian | Y | Y | Y | | Y | Y |
| 6196 | SAMN15879252 | T1D ICI | 20-30 | Female | 26.6 | 0.48 | 15 | GADA+ mIAA+ | African Am | Y | Y | Y | | | |
| 6198 | SAMN15879254 | T1D ICI | 20-30 | Female | 23.1 | nd | 3 | GADA+ IA2A+ mIAA+ ZnT8A+ | Hispanic | Y | Y | | | Y | |
| 6209 | SAMN15879265 | T1D ICI | 0-10 | Female | 15.9 | 0.1 | 0.25 | IA2A+ mIAA+ ZnT8A+ | Caucasian | Y | Y | Y | Y | Y | |
| 6211 | SAMN15879267 | T1D ICI | 20-30 | Female | 24.4 | nd | 4 | GADA+ IA2A+ mIAA+ ZnT8A+ | African Am | Y | Y | Y | | Y | Y |
| 6212 | SAMN15879268 | T1D ICI | 20-30 | Male | 29.1 | nd | 5 | mIAA+ | Caucasian | Y | | | | Y | Y |

| | | | | | | | | | | | | | | | |
|---|---|---|---|---|---|---|---|---|---|---|---|---|---|---|---|
| 6228 | SAMN15879284 | T1D ICI | 10-20 | Male | 17.4 | 0.1 | 0 | GADA+ IA2A+ ZnT8A+ | Caucasian | Y | | | Y | Y | |
| 6243 | SAMN15879299 | T1D ICI | 10-20 | Male | 21.3 | 0.42 | 5 | mIAA+ | Caucasian | | | | | Y | Y |
| 6245 | SAMN15879301 | T1D ICI | 20-30 | Male | 23.2 | nd | 7 | GADA+ IA2A+ | Caucasian | Y | | | | Y | |
| 6247 | SAMN15879303 | T1D ICI | 20-30 | Male | 24.3 | 0.47 | 0.6 | mIAA+ | Caucasian | Y | Y | Y | Y | | |
| 6264 | SAMN15879318 | T1D ICI | 10-20 | Female | 22 | nd | 9 | Negative | Caucasian | Y | | | | | |
| 6265 | SAMN15879319 | T1D ICI | 10-20 | Male | 12.9 | 0.06 | 8 | GADA+ mIAA+ | Caucasian | Y | | | | | Y |
| 6268 | SAMN15879322 | T1D ICI | 10-20 | Female | 26.6 | 0.05 | 3 | mIAA+ | Caucasian | | | | | | Y |
| 6302 | SAMN15879356 | T1D ICI | 30-40 | Male | 20.5 | 0.17 | 32.5 | Negative | African Am | Y | | | | | |
| 6306 | SAMN15879360 | T1D ICI | 10-20 | Male | 24.5 | nd | 5 | mIAA+ | Caucasian | Y | | | | | Y |
| 6307 | SAMN15879361 | T1D ICI | 40-50 | Female | 19.5 | nd | 10 | GADA+ mIAA+ | Caucasian | Y | | | | | |
| 6325 | SAMN15879379 | T1D ICI | 20-30 | Female | 31.2 | 0.14 | 6 | GADA+ IA2A+ mIAA+ | African Am | Y | | | | | Y |
| 6328 | SAMN15879382 | T1D ICI | 30-40 | Male | 24 | nd | 20 | GADA+ mIAA+ | Hispanic | Y | | | | | |
| 6337 | SAMN15879391 | T1D ICI | 20-30 | Female | 17.9 | nd | 5 | mIAA+ | Caucasian | Y | | | | | |
| 6342 | SAMN15879396 | T1D ICI | 10-20 | Female | 24.3 | 0.26 | 2 | IA2A+ mIAA+ | Caucasian | Y | Y | Y | | | Y |
| 6362 | SAMN15879415 | T1D ICI | 20-30 | Male | 28.5 | 0.38 | 0 | GADA+ | Caucasian | Y | Y | | | | Y |
| 6367 | SAMN15879420 | T1D ICI | 20-30 | Male | 25.7 | 0.39 | 2 | Negative | Caucasian | Y | Y | | | | Y |
| 6371 | SAMN15879424 | T1D ICI | 10-20 | Female | 16.6 | 0.11 | 2 | GADA+ IA2A+ mIAA+ ZnT8A+ | Caucasian | Y | Y | | | | |
| 6380 | SAMN15879433 | T1D ICI | 10-20 | Female | 14.6 | 0.22 | 0 | Negative | African Am | Y | Y | | | | |
| 6405 | SAMN15879458 | T1D ICI | 20-30 | Female | 42.5 | 1.84 | 0.6 | GADA+ IA2A+ ZnT8A+ | Hispanic | Y | Y | | | | |
| 6026 | SAMN15879083 | T1D IDI | 20-30 | Male | 24.1 | nd | 9 | mIAA+ | Caucasian | Y | | | | Y | |
| 6031 | SAMN15879088 | T1D IDI | 30-40 | Male | 24.5 | nd | 35 | mIAA+ | Caucasian | Y | | | | | |
| 6035 | SAMN15879092 | T1D IDI | 30-40 | Male | 27.1 | nd | 28 | mIAA+ | Caucasian | Y | Y | | | | |
| 6039 | SAMN15879096 | T1D IDI | 20-30 | Female | 23.4 | nd | 12 | GADA+ IA2A+ mIAA+ ZnT8A+ | Caucasian | Y | | | | Y | |
| 6041 | SAMN15879098 | T1D IDI | 20-30 | Male | 28.4 | nd | 23 | Negative | Caucasian | Y | | | | | |
| 6045 | SAMN15879102 | T1D IDI | 20-30 | Male | 23.1 | nd | 8 | mIAA+ ZnT8A+ | Caucasian | Y | | | | Y | |
| 6063 | SAMN15879120 | T1D IDI | 0-10 | Male | 23.8 | nd | 3 | mIAA+ | Caucasian | Y | Y | Y | | Y | Y |
| 6066 | SAMN15879123 | T1D IDI | 70-80 | Male | 30.9 | nd | 74 | IA2A+ mIAA+ | Caucasian | Y | | | | | |
| 6067 | SAMN15879124 | T1D IDI | 30-40 | Female | 26.8 | nd | 8 | Negative | Hispanic | Y | | | | | |

| | | | | | | | | | | | | | | | |
|---|---|---|---|---|---|---|---|---|---|---|---|---|---|---|---|
| 6076 | SAMN15879133 | T1D IDI | 20-30 | Male | 18.8 | nd | 15 | GADA+ mIAA+ | Caucasian | Y | | | | Y | |
| 6077 | SAMN15879134 | T1D IDI | 30-40 | Female | 22 | nd | 19 | mIAA+ | Caucasian | Y | | | | Y | |
| 6079 | SAMN15879136 | T1D IDI | 10-20 | Female | 18.6 | nd | 8 | Negative | Caucasian | Y | | | | | Y |
| 6083 | SAMN15879140 | T1D IDI | 10-20 | Female | 18.4 | nd | 11 | mIAA+ | Caucasian | Y | Y | Y | | | |
| 6087 | SAMN15879144 | T1D IDI | 10-20 | Male | 21.9 | nd | 4 | mIAA+ ZnT8A+ | Caucasian | Y | Y | Y | | | |
| 6089 | SAMN15879146 | T1D IDI | 10-20 | Male | 26 | nd | 8 | mIAA+ | Caucasian | Y | Y | Y | | | |
| 6119 | SAMN15879176 | T1D IDI | 0-10 | Male | 19.4 | nd | 14 | GADA+ mIAA+ | Caucasian | Y | | | | Y | Y |
| 6128 | SAMN15879185 | T1D IDI | 30-40 | Female | 22.2 | nd | 31.5 | mIAA+ | Caucasian | Y | Y | Y | | Y | Y |
| 6135 | SAMN15879192 | T1D IDI | 40-50 | Male | 28.7 | nd | 21 | GADA+ mIAA+ | Caucasian | Y | Y | Y | | | |
| 6138 | SAMN15879195 | T1D IDI | 40-50 | Female | 33.7 | nd | 41 | mIAA+ | Caucasian | Y | Y | Y | | | |
| 6141 | SAMN15879198 | T1D IDI | 30-40 | Male | 26 | nd | 28 | GADA+ IA2A+ mIAA+ ZnT8A+ | Caucasian | Y | Y | Y | | Y | |
| 6143 | SAMN15879200 | T1D IDI | 30-40 | Female | 26.1 | nd | 7 | IA2A+ mIAA+ | Caucasian | Y | Y | Y | | Y | Y |
| 6145 | SAMN15879202 | T1D IDI | 10-20 | Male | 23.1 | 0.06 | 11 | GADA+ mIAA+ ZnT8A+ | Caucasian | Y | Y | Y | | | |
| 6148 | SAMN15879204 | T1D IDI | 10-20 | Male | 23.9 | nd | 7 | GADA+ mIAA+ | Caucasian | Y | Y | Y | | | Y |
| 6152 | SAMN15879208 | T1D IDI | 20-30 | Female | 30.1 | nd | 12 | ZnT8A+ | Caucasian | Y | Y | Y | | | |
| 6155 | SAMN15879211 | T1D IDI | 50-60 | Female | 26 | nd | 43 | mIAA+ | Caucasian | Y | Y | Y | | | |
| 6159 | SAMN15879215 | T1D IDI | 50-60 | Female | 35.5 | nd | 44 | mIAA+ | Caucasian | Y | Y | Y | | | |
| 6161 | SAMN15879217 | T1D IDI | 10-20 | Female | 36.1 | nd | 7 | IA2A+ mIAA+ | Caucasian | Y | Y | Y | | | Y |
| 6163 | SAMN15879219 | T1D IDI | 30-40 | Male | 25.5 | nd | 30 | IA2A+ mIAA+ | Caucasian | Y | Y | Y | | | |
| 6169 | SAMN15879225 | T1D IDI | 20-30 | Female | 25 | nd | 15 | GADA+ mIAA+ | Hispanic | Y | Y | Y | | | |
| 6173 | SAMN15879229 | T1D IDI | 40-50 | Male | 23.9 | nd | 15 | Negative | Caucasian | Y | Y | Y | | | |
| 6205 | SAMN15879261 | T1D ICI | 40-50 | Female | 22.6 | 0.14 | 33 | mIAA+ | Caucasian | Y | Y | Y | | | |
| 6207 | SAMN15879263 | T1D IDI | 10-20 | Female | 24.4 | nd | 10 | IA2A+ mIAA+ ZnT8A+ | African Am | Y | Y | Y | | | |
| 6208 | SAMN15879264 | T1D IDI | 30-40 | Female | 23.4 | nd | 16 | Negative | Caucasian | Y | Y | Y | | | |
| 6224 | SAMN15879280 | T1D IDI | 20-30 | Female | 22.8 | nd | 1.5 | Negative | Caucasian | | | | Y | | Y |
| 6324 | SAMN15879378 | T1D IDI | 20-30 | Male | 26.2 | nd | 2 | GADA+ mIAA+ | Hispanic | Y | Y | Y | Y | | |
| 6418 | SAMN15879471 | T1D IDI | 20-30 | Male | 26.4 | nd | 11 | GADA+, IA-2A+, mIAA+*, ZnT8+ | Caucasian | | | | | | Y |

**ESM Data 1.** Description of the positive control experiment

**Description of the preparation of the positive control samples for UCL RNA-Seq studies**

An EV-negative pancreas sample of a non-diabetic organ donor from the PanFin study [Tauriainen et al 2010] was homogenized using a Silent Crusher S homogenizer (Heidolph, Schwabach, Germany). The pancreas extract was divided into aliquots and spiked with infectious virus preparations of coxsackievirus B1 (CBV1). The virus was propagated in GMK (green monkey kidney) cells, and cell culture supernatant was used as virus source. Pancreas samples were spiked with virus dilutions ($10^{-3}$, $10^{-6}$, $10^{-7}$, $10^{-8}$, $10^{-9}$) and immediately frozen at −80 °C. Schematic presentation of the CVB1 spiked in pancreas dilution series used as positive control experiment for RNA-Seq analyses in UCL is presented below:

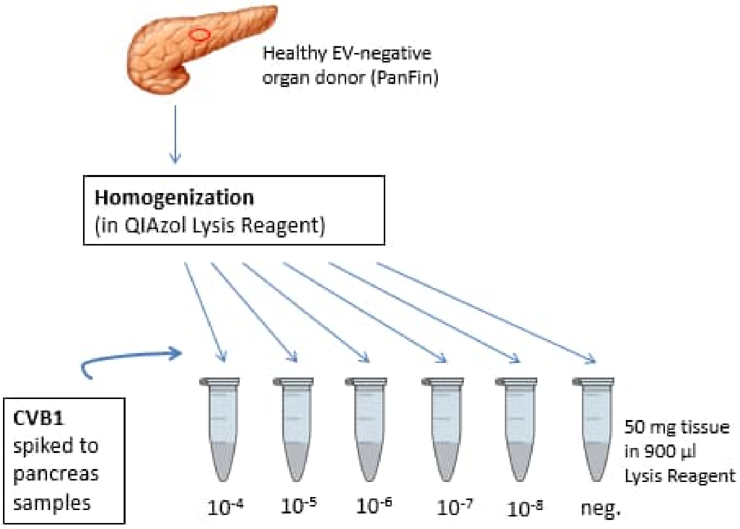


Ref. Tauriainen, S.; Salmela, K.; Rantala, I.; Knip, M.; Hyöty, H. Collecting high-quality pancreatic tissue for experimental study from organ donors with signs of beta-cell autoimmunity. *Diabetes Metab. Res. Rev.* **2010**, *26*, 585–592.

**ESM Table 2:** Detection of CVB1 specific sequences by metagenomic sequencing in homogenized pancreas samples spiked with different dilutions with CVB1 (five spiked-in samples with CVB1 and one negative un-spiked control sample).

| | Raw | QC | QC | CVB1 aligned | CVB1 |
|---|---|---|---|---|---|
| spike-in dilution of CVB1 | # pairs | # pairs | % | # reads | % |
| 0 (negative control) | 4728119 | 4349094 | 0.92 | 2reads->1 pair | |
| 10e-8 | 4223286 | 3971535 | 0.94 | 32 | 4.0E-006 |
| 10e-7 | 3710861 | 3351818 | 0.90 | 283 | 4.2E-005 |
| 10e-6 | 3546996 | 3229128 | 0.91 | 2595 | 4.0E-004 |
| 10e-5 | 4392146 | 4015026 | 0.91 | 12237 | 1.5E-003 |
| 10e-4 | 2448108 | 2149775 | 0.88 | 132770 | 3.1E-002 |

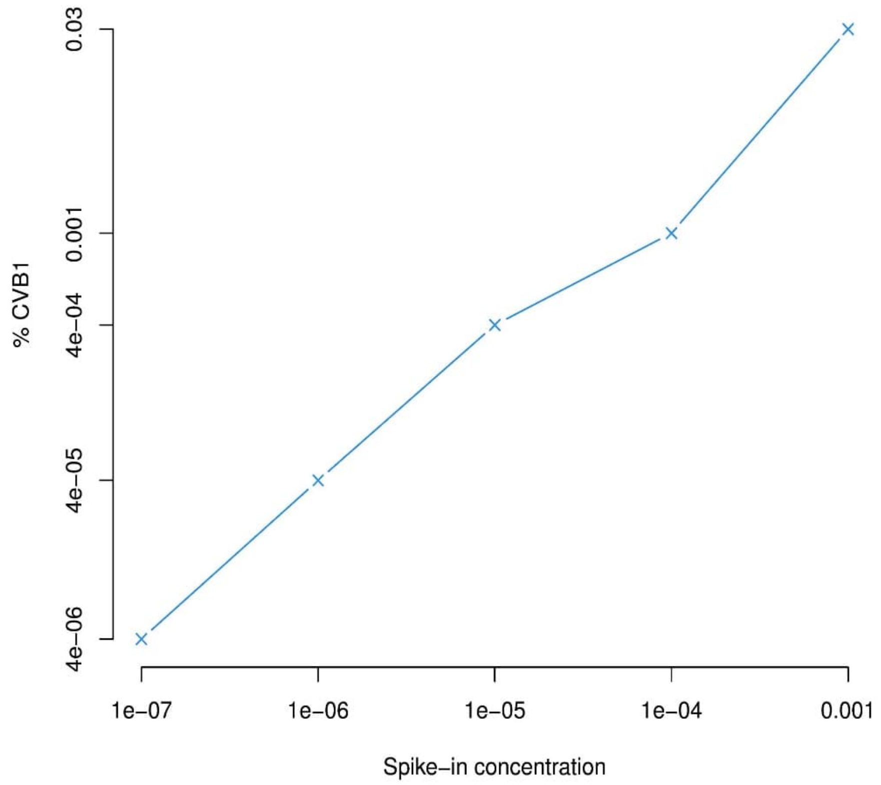


**ESM. Fig. 1.** Linear relationship between CVB1 spike-in concentrations and % on the target viral reads. Both axes are in log10 scale.

**ESM Table 3**. UCL Step I RNA-seq analyses from nPOD samples: Taxonomic classification of the "microbial" reads with metaMix resulted in a similar profile for all samples, showing no difference between case and control donors. The majority of reads was assigned to Enterobacteria phage phiX174, the positive control for Illumina sequencing. The rest of the reads were divided between various environmental bacteria and the “unknown” bin. Table represents, as an example, a general profile and relative abundances in 6070 (T1D ICI), 6098 (ND), 6141 (T1D IDI).

| Organisms | mean abundance % (sd) |
|---|---|
| *Enterobacteria phage phiX174* | 62 (0.03) |
| *Environmental bacteria* | 25 (0.02) |
| *Unknown* | 13 (0.01) |

T1D ICI – type 1 diabetes with insulin containing islets ; T1D IDI – type 1 diabetes with isulin deficient islets ; ND – non-diabetic

**ESM Table 4:** Example of metaMix summary profile for one case, sequenced using the sequence capture approach to enrich enterovirus specific sequences (Step III).

| taxon id | scientific name | assigned reads | posterior prob |
|---|---|---|---|
| unknown | unknown | 188927 | 1 |
| 9606 | *Homo sapiens* | 2745 | 1 |
| 374840 | *Enterobacteria phage phiX174 sensu lato* | 698 | 0.92 |

**ESM Table 5.** Group comparisons of enterovirus positivity in the pancreas by RT-PCR (relates to **Fig.2 A**)

| Donor group comparisons | P value (Fisher exact test 2-sided) | P value corrected (10 comparisons) |
|---|---|---|
| **ND vs Aab+** | **0.0004** | **0.004** |
| ND vs Aab++ | 1.000 | N.S. |
| ND vs T1D-ICI | 0.4713 | N.S. |
| ND vs T1D-IDI | 0.1405 | N.S. |
| Aab+ vs Aab++ | 0.0225 | N.S. |
| Aab+ vs T1D-ICI | **0.0131** | N.S. |
| **Aab+ vs T1D-IDI** | **0.000** | **0.000** |
| Aab++ vs T1D-ICI | 0.5628 | N.S. |
| Aab++ vs T1D-IDI | 1.000 | N.S. |
| **T1D-ICI vs T1D-IDI** | **0.0225** | N.S. |

**ESM Table 6**. Pancreas enterovirus positivity with regards to active islet autoimmunity (IA) (relates to **Fig.2 B**). Significant p-values are bolded.

| Donor group comparisons | P value (Fisher exact test 2-sided) | P value corrected (6 comparisons) |
|---|---|---|
| ND vs IA and ICIs | **0.0078** | **0.0468** |
| ND vs Autoimmunity | **0.0639** | N.S. |
| Autoimmunity vs Autoimmunity and ICIs | 0.367 | N.S. |
| Autoimmunity and ICIs vs No autoimmunity | **0.001** | **0.006** |
| ND vs No autoimmunity | 0.2907 | N.S. |
| Autoimmunity vs No autoimmunity | **0.004** | **0.04** |

**ESM Table 7**. Enterovirus detection in the spleen **by RT-PCR**. Corrected p-values for multiple comparisons (N=10) are also shown (relates to **Fig. 3A**)

| Donor group comparisons | P value (Fisher exact 2-sided) | P value corrected (10 comparisons) |
|---|---|---|
| ND vs Aab+ | 0.5597 | N.S. |
| ND vs Aab++ | 1 | N.S. |
| ND vs T1D ICI | 0.6823 | N.S. |
| ND vs T1D IDI | 0.6392 | N.S. |
| Aab+ vs Aab++ | 0.3684 | N.S. |
| Aab+ vs T1D ICI | 0.2645 | N.S. |
| Aab+ vs T1D IDI | 1 | N.S. |
| Aab++ vs T1D ICI | 1 | N.S. |
| Aab++ vs T1D IDI | 0.4184 | N.S. |
| T1D ICI vs T1D IDI | 0.3129 | N.S. |

**ESM Table 8**. RNA quality numbers (RQN) for selected nPOD donors in different tissues vs. enterovirus positivity by RT-PCR.

| | | Pancreas | | Spleen | | Duodenum | |
|---|---|---|---|---|---|---|---|
| **Donor ID** | **Donor type** | **EV PCR** | **RQN** | **EV PCR** | **RQN** | **EV PCR** | **RQN** |
| **6097** | **ND** | | | POS | 6.2 | NEG | 2.2 |
| **6044** | **Aab+** | POS | 1 | | | | 2 |
| **6046** | **T1D ICI** | POS | 5.3 | POS | 1.1 | | |
| **6087** | **T1D IDI** | | | POS | 2.4 | | |
| **6090** | **Aab+** | NEG | 6.3 | | | | |
| **6101** | **Aab+** | POS | 2 | | | | |
| **6102** | **ND** | | | | | NEG | 1.5 |
| **6106** | **ND** | | | | | NEG | 1 |
| **6112** | **ND** | | | POS | 1 | NEG | 1 |
| **6123** | **Aab+** | POS | 2.8 | | | | |
| **6154** | **Aab+** | POS | 3 | | | | |
| **6156** | **Aab+** | POS | 5 | | | | |
| **6158** | **Aab++** | POS | 3.9 | | | | |
| **6167** | **Aab++** | NEG | 1.5 | | | | |
| **6209** | **T1D ICI** | POS | 5.7 | POS | 1.6 | NEG | 1.3 |

| | | | | | | | |
|---|---|---|---|---|---|---|---|
| **6247** | **T1D ICI** | NEG | 7.8 | | | NEG | 4.4 |
| **6267** | **Aab++** | NEG | 5.8 | | | NEG | 3.6 |
| **6324** | **T1D IDI** | | | | | NEG | 7.3 |
| **6342** | **T1D ICI** | | | | | NEG | 3 |

**ESM Table 9**. Enterovirus detection in the spleen by enterovirus propagation. Significant p-values are bolded. Corrected p-values for multiple comparisons (N=10) are also shown (Relates to **Fig. 2B**)

| **Donor group comparisons** | **P value (Fisher exact 2-sided)** | **P value corrected*** |
|---|---|---|
| ND vs Aab+ | 1 | N.S |
| ND vs Aab++ | 1 | N.S |
| ND vs T1D ICI | **0.0069** | N.S |
| ND vs T1D IDI | **0.001** | **0.01** |
| Aab+ vs Aab++ | 1 | N.S |
| Aab+ vs T1D ICI | 0.1619 | N.S |
| Aab+ vs T1D IDI | 0.0406 | N.S |
| Aab++ vs T1D ICI | 0.4706 | N.S |
| Aab++ vs T1D IDI | 0.1091 | N.S |
| T1D ICI vs T1D IDI | 0.4013 | N.S |
